# Spatial-temporal and phylogenetic analyses of epidemiologic data to help understand the modes of transmission of endemic typhoid fever in Samoa

**DOI:** 10.1101/2022.03.28.22272797

**Authors:** Michael J. Sikorski, Jianguo Ma, Tracy H. Hazen, Sachin N. Desai, Siaosi Tupua, Susana Nimarota-Brown, Michelle Sialeipata, Savitra Rambocus, Susan A. Ballard, Mary Valcanis, Robert E. Thomsen, Roy M. Robins-Browne, Benjamin P. Howden, Take K. Naseri, Myron M. Levine, David A. Rasko

## Abstract

*Salmonella enterica* serovar Typhi (*S*. Typhi) is either widely distributed or proximally transmitted via fecally-contaminated food or water to cause typhoid fever. In Samoa, where endemic typhoid fever has persisted over decades despite water quality and sanitation improvements, the local patterns of *S*. Typhi circulation remain undistinguished. From April 2018-June 2020, epidemiologic data and GPS coordinates were collected during household investigations of 260 acute cases of typhoid fever, and 27 asymptomatic shedders of *S*. Typhi were detected among household contacts. Spatial and temporal distributions of cases were examined using Average Nearest Neighbor and space-time hotspot analyses. In rural regions, infections occurred in sporadic, focal clusters contrasting with persistent, less clustered cases in the Apia Urban Area. Restrictions to population movement during nationwide lockdowns in 2019-2020 were associated with marked reductions of cases. Phylogenetic analyses of isolates with whole genome sequences (n=186) revealed one dominant genotype 3.5.4 (n=181/186) that contains three Samoa-exclusive sub-lineages: 3.5.4.1, 3.5.4.2, and 3.5.4.3. Variables of patient sex, age, and geographic region were examined by phylogenetic groupings, and significant differences (p<0.05) associated genetically-similar isolates in urban areas with working ages (20-49 year olds), and in rural areas with age groups typically at home (<5, 50+). Isolates from asymptomatic shedders were among all three sub-lineages. Whole genome sequencing also corroborated bacterial genetic similarity in 10/12 putative epidemiologic linkages among cases and asymptomatic shedders as well as 3/3 repeat positives (presumed relapses), with a median of one single nucleotide polymorphism difference. These findings highlight various patterns of typhoid transmission in Samoa that differ between urban and rural regions as well as genomic subtypes. Asymptomatic shedders, detectable only through household investigations, are likely an important reservoir and mobile agent of infection. This study advances a “Samoan *S*. Typhi framework” that supports current and future typhoid surveillance and control efforts in Samoa.

**AUTHOR SUMMARY:** Many typhoid endemic countries have evident transmission contributions from widely distributed contaminated water supplies and/or asymptomatic *S*. Typhi carriers who intermittently cause sporadic outbreaks. However, these patterns have not yet been examined in the island nation of Samoa, where typhoid has remained endemic for decades. In this study, we incorporated the discerning powers of spatial-temporal cluster analyses as well as phylogenetics of whole genome sequences (WGS) of *S*. Typhi isolates to examine detailed epidemiologic data collected through household investigations of culture-confirmed cases of typhoid fever occurring in Samoa from April 2018 through June 2020. We detected patterns consistent with both modes of transmission, varying between urban and rural regions, and we provided evidence of intra-household transmission of genetically similar isolates, thereby supporting a majority of putative epidemiologic linkages made during household investigations, and identifying important roles for asymptomatic shedders of *S*. Typhi. These findings advance our understanding of persistently endemic typhoid fever in Samoa and directly support the efforts of the Samoa Typhoid Fever Control Program of the Ministry of Health of Samoa.

## INTRODUCTION

Typhoid fever is a potentially fatal human host-restricted infection caused by ingestion of *Salmonella enterica* serovar Typhi (*S*. Typhi) via contaminated food or water. Typhoid incidence, reported as cases per 100,000 people per year, is stratified into low (<10), moderate (10-100), high (>100–<500), and very high (≥500), with different transmission mechanisms capable of maintaining typhoid fever at different incidence levels [1]. Widespread distribution of fecally-contaminated indirect sources (called “vehicles”), such as water supplies [2,3], crops irrigated with raw sewage [4,5], or processed foods [6,7], results in amplified, high and very high incidence transmission of *S*. Typhi. Sporadic, focal outbreaks of typhoid fever are typically associated with asymptomatic shedding of *S*. Typhi from a temporary (weeks to months) [8] or chronic (lifelong) carrier, such as “Typhoid Mary” [9]. These asymptomatic shedders inadvertently contaminate food that is consumed by contacts in close proximity, such as household members or attendees at gatherings (e.g. picnics and celebrations) [10].

Samoa, an island nation in the Polynesian sub-region of Oceania (the continental grouping of thousands of islands comprising 14 countries and 9 dependencies in the Central and South Pacific Ocean), has endured persistent endemic typhoid fever for decades despite improvements in drinking water quality, sanitation infrastructure, and economic development [11–14]. We have previously described endemic typhoid in Samoa by person, place, and time using blood culture surveillance data from 2008-2019 [14] and characterized the population structure and evolutionary origins of a collection of *S*. Typhi genomes from Samoa spanning 1983-2020 [15]. However, it remains unclear to what extent endemic typhoid in Samoa is sustained by widespread, continuous distribution of an unknown fecally-contaminated vehicle (e.g., municipal water supplies) or focal, sporadic clusters caused by ingestion of food contaminated by temporary or chronic shedders. Differing in spatial and temporal distribution, these transmission cycles can be examined using geospatial point-pattern analyses [16,17]. Additionally, when paired with data from epidemiologic investigations, whole genome sequencing (WGS) analyses can correlate epidemiology with phylogeny, infer patterns of infection by genetic subtype, and quantify relatedness between isolates to evaluate suspected transmission events [18].

In Samoa, teams from the Samoa Typhoid Fever Control Program (STFCP) of the Ministry of Health (MoH) perform epidemiological investigations of the households of every culture-confirmed typhoid case to ascertain clinical status, environmental risks, prevalence of *S*. Typhi among household contacts, and evidence of epidemiologic association to other cases. In this study, we combined epidemiologic data collected through household investigations from April 2018 through June 2020 with geospatial point-pattern statistical tools and the high resolution discernment of WGS. We examined the spatial and temporal distribution of typhoid cases, as well as the sex, age, and geographic occurrences of infections by phylogenetic groupings. Putative epidemiologic linkages identified through case investigations and rare repeat positive isolates from the same individual were examined for genomic similarity and provided primary data to inform a phylogenetic threshold metric of relatedness among endemic *S*. Typhi strains. Our findings can help elucidate how *S*. Typhi circulates in Samoa and support the further integration of WGS into the epidemiologic surveillance and control efforts of typhoid control programs.

## METHODS

This study received ethical clearance from the Health Research Committee of the MoH of Samoa, as well as The University of Maryland, Baltimore (UMB) Institutional Review Board protocol HP-00087489.

### Study setting

This study took place from 2018-2020 in Samoa, a Pacific Island nation of area 2,842 km^2^ (1,097 sq. mi) and ∼200,000 population. Samoa comprises two populated islands, Upolu and Savaii, each with a central clinical laboratory and several peripheral health facilities (Fig 1). Typhoid fever is a notifiable infectious disease in Samoa that triggers epidemiologic investigation of any and all culture-confirmed cases [19,20]. Since 2018, the STFCP has strengthened epidemiologic surveillance for typhoid fever, improved clinical and environmental microbiology laboratory capacity, and initiated WGS at a collaborating regional reference laboratory in Melbourne, Australia. Typhoid Fever Epidemiologic “SWAT” Teams (equipped with specialized epidemiologic tools and tactics), one on each island, were trained to perform epidemiologic investigations of the household (and/or workplace or school) of every bacteriologically confirmed case of typhoid fever occurring anywhere in Samoa.

**Fig 1.**
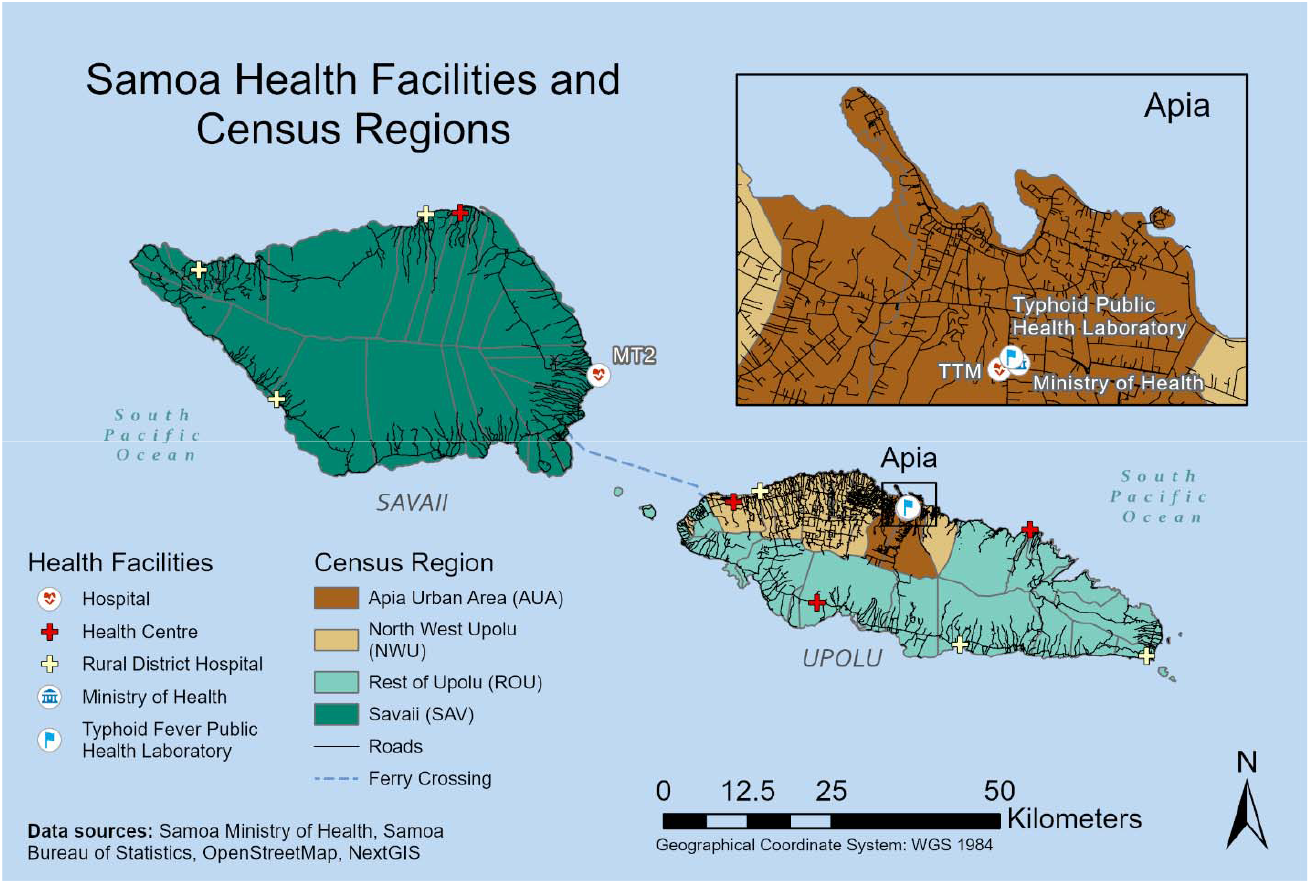
Map of Samoa with census regions, roads, and health facilities. Samoa is comprised of two primary islands, Upolu to the east and Savaii to the west. About three quarters of the population and the capital town of Apia are located on Upolu. The black lines indicate roads associated with populated areas, which are predominantly located around the periphery of the islands except for the urban areas where inland settlements are increasing. The islands are subdivided into four statistical census regions, 43 political districts (outlined in light grey), and >300 villages (not depicted). The four colored geographic areas differentiate the four census regions: Apia Urban Area (AUA), North West Upolu (NWU), Rest of Upolu (ROU), and Savaii (SAV). AUA is most dense with ∼25% of the total population. Apia is the capital seat of government and includes the Ministry of Health headquarters, Samoa Typhoid Fever Public Health Laboratory, and Tupua Tamasese Meaole (TTM) national hospital, which processes the blood cultures for Upolu (insert). NWU is mixed peri-urban and rural in population density and separates Apia from the inter-island ferry (navy dashed line) and adjacent international airport (not shown). The Malietoa Tanumafili II (MT2) hospital processes blood cultures for Savaii. The health facility icons indicate the two central hospitals with clinical microbiology laboratories, TTM and MT2, and the 6 district hospitals and 4 peripheral health centers.

### Case ascertainment

Febrile patients who presented to an MoH health facility with clinical signs and symptoms of typhoid fever [21] triggered the collection of a blood culture. Patients from whom *S*. Typhi was isolated and confirmed by standard methods [22,23] were considered acute blood culture-confirmed cases of typhoid fever. Every such case was reported to the Typhoid SWAT Team on the incident island for expeditious investigation. On three occasions, the same individual presented to a health facility with culture-confirmed typhoid fever within ∼1 month of a prior confirmed typhoid infection. These three repeat positive cases (presumed relapses) were counted as distinct cases but considered epidemiologically linked by person.

### Case investigations and epidemiologic linkages

During household visits, the Typhoid SWAT teams evaluated environmental risk factors for typhoid transmission and ascertained the clinical wellbeing of the index case and their household contacts. Global positioning system (GPS) coordinates of the home were collected using Garmin GPSMAP 64sc handheld navigators. The Typhoid SWAT teams also sought evidence of epidemiologic links to other recent typhoid cases occurring within two months in the same home/venue, in neighboring homes, or in contacts living elsewhere in Samoa. To detect temporary or chronic asymptomatic shedders of *S*. Typhi among household contacts, up to three rectal swabs collected 1-2 days apart from each contact were transported in Cary-Blair transport medium to the Typhoid Fever Public Health Laboratory in Apia and examined using standard bacteriologic methods [22,23]. Asymptomatic contacts from whom *S*. Typhi was isolated from stool cultures were referred for treatment [24]. These asymptomatic shedders were considered epidemiologically linked to the index case by person, place, and time.

### Spatial and temporal point pattern analyses

Using the spatial statistics toolbox in Esri ArcGIS Pro v2.9 [25], we performed two point pattern cluster analyses of case households (n=260) to ascertain significant (p<0.05) spatial and spatial-temporal clusters. First, we used the Average Nearest Neighbor (ANN) analysis to determine if the spatial distribution of points (i.e., case households) was clustered based on the average distances between them [26,27]. The resulting nearest neighbor ratio (R_n_) ranged from zero (significant clustering) to 2.15 (uniform equidistant dispersion) and was supported by critical values (Z-score and p-value); an R_n_ equal to 1 indicated randomness or no detectable spatial pattern (S1 and S2 Figs). The ANN analysis was performed on household coordinates in aggregate and then by each of the four census regions (Fig 1, S3A Fig), which separate the two Samoan islands into the Apia Urban Area (AUA; urban; ∼625 persons per km^2^), North West Upolu (NWU; mixed peri-urban and rural; ∼275 persons per km^2^), Rest of Upolu (ROU; rural; ∼60 persons per km^2^) and Savaii (SAV; rural; ∼26 persons per km^2^) [28].

We also used the Space-Time Hotspot analysis tool to detect significant clustering of acute case households over time. Case household coordinates were binned into 1 km^2^ x 2-month space-time cubes and visualized in three dimensions with red shading to indicate space-time clustering confidence levels (Fig 2). Detailed descriptions and methodologies for the ANN and Space-Time Hotspot tools are included in the Supplementary Information.

**Fig 2.**
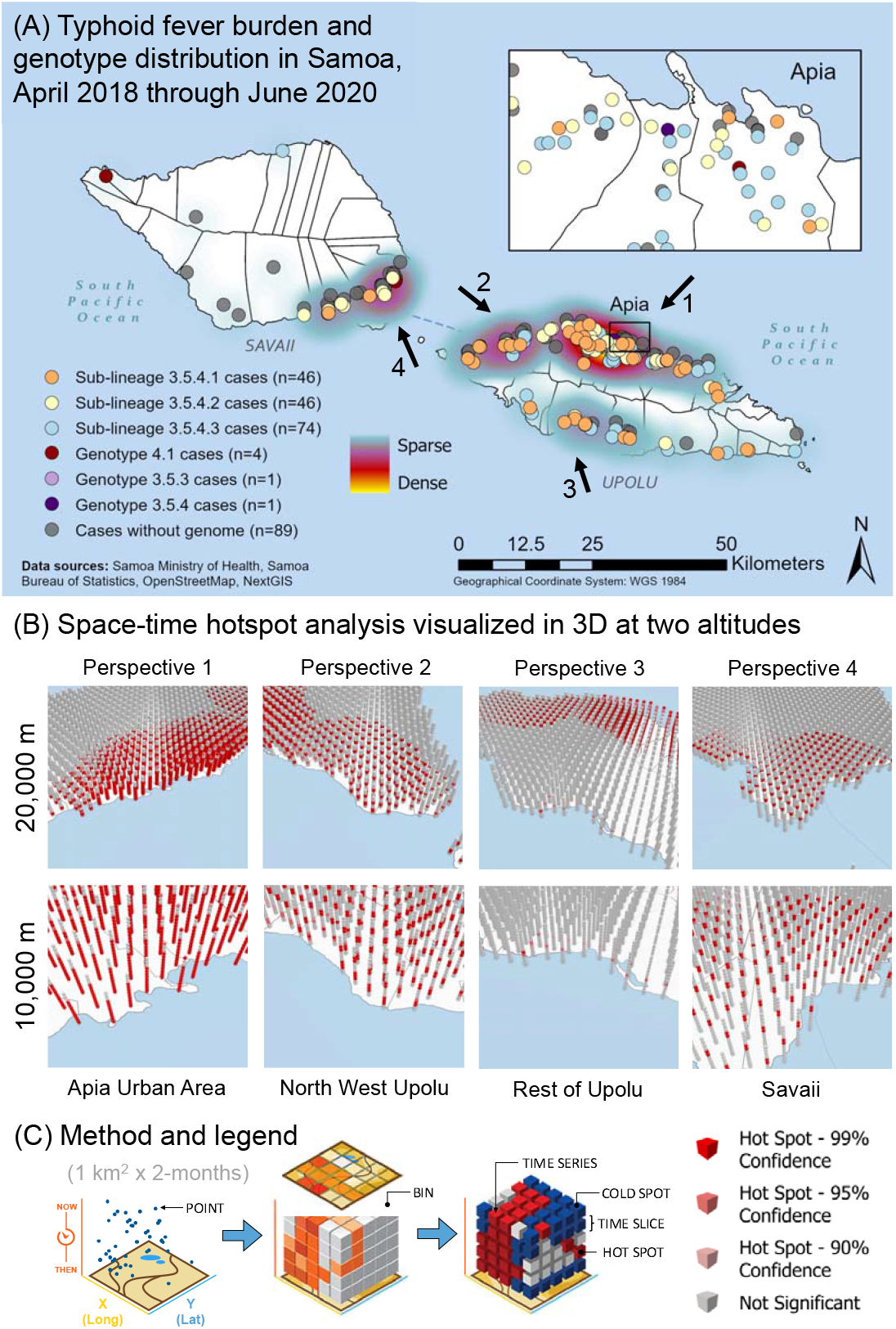
Spatial-temporal and genomic analysis of typhoid fever cases (burden) in Samoa from April 2018 through June 2020. **(A)** Using ArcGIS Pro v2.9, household GPS coordinates of acute cases of *S*. Typhi (n=260) from April 27, 2018 to June 9, 2020 are plotted on a map of Samoa and colored according to *S*. Typhi genotype/sub-lineage. A point density heatmap is inferred from case coordinates. The four prominent hotspots are identified with arrows labeled 1-4, which also indicate aerial perspectives shown in the reconstructions in panel B. **(B)** Coordinates were binned into space-time cubes of 1km^2^ x 2-months (see panel C and Supplementary Information for more details) and tested for significant clustering by the emerging hotspot analysis implemented in ArcGIS Pro. The resulting 3D space-time hotspots were visualized from an aerial view of 20,000 meters and 10,000 meters focusing on the four typhoid hotspots identified in panel A. In the 3D renderings, each vertical column on the map represents a stack of cubes over 1 km^2^ area and with a height representing 2 months of time, totaling 13 time steps, or all 26 months of this study. Aggregation of points in both space and time generate a space-time hotspot, visible as a red cube or band of several adjacent cubes. The intensity of red shading indicates statistical confidence. **(C)** To generate space-time cubes, points were graphed in a 3-dimensional plane based on coordinates and time, and then binned into cubes of space and time dimensions. Counts within each cube were compared to nearby cubes to extrapolate hot and cold spots with statistical support (99%, 95%, 90%, not significant). Hotspots in the x-y plane occur within a time slice. Hotspots inferred from the z-direction fall within a time series. Adapted with permission from Esri (Redlands, CA: Environmental Systems Research Institute, Inc.).

### Whole genome sequencing and phylogenetic analysis

All available *S*. Typhi blood isolates from acute cases and stool isolates from asymptomatic shedders were shipped to the Microbiological Diagnostic Unit Public Health Laboratory (University of Melbourne, Australia) for whole genome sequencing, as previously described [15]. Briefly, extracted DNA was sequenced on the Illumina NextSeq 500 with 150-cycle paired end chemistry. Genotypes were determined from reads mapped to the reference strain CT18 (GCA_000195995.1) using GenoTyphi v1.9.1 [29,30]. Genotype 3.5.4.1, 3.5.4.2, and 3.5.4.3 sub-lineages unique to Samoa were assigned based on canonical single nucleotide polymorphisms (SNPs) against CT18, as previously described [15]. To generate a locally referenced maximum-likelihood (ML) phylogeny, reads were assembled and core genome SNPs were identified relative to the 2012 Samoan *S*. Typhi reference strain H12ESR00394-001A (GCA_001118185.2). After removal of recombination sites, a ML phylogeny was inferred using the generalized time-reversible (GTR) site substitution model with a Gamma rate distribution and the Lewis ascertainment bias correction (ASC_GTRGAMMA) and 100 bootstrap pseudo-replicates in RAxML v8.2.10 [31]. Pairwise SNP distances between genomes were calculated using snp-dists v0.8.2 [32] and compared among epidemiologically linked isolates. Detailed protocols are found in the Supplementary Information, with accession numbers listed in S1 Table and assemblies available on Figshare (doi: 10.6084/m9.figshare.18665686).

### Epidemiologic analysis of genotypes/sub-lineages

Patient characteristics (sex, age group, and mean age) were assessed for differences between all cases and the subset of sequenced isolates using Pearson’s chi-squared test of independence for the categorical variables and unpaired two-tailed T test of means for mean age. Phylogenetic groupings of *S*. Typhi were examined for associations with epidemiologic variables (infection status, sex, age group, census region, year) using Fisher’s exact test. The ANN statistic was performed in ArcGIS Pro v2.9 on groupings of household coordinates by each genotype 3.5.4 sub-lineage (S3B-D Fig). The absolute and relative counts of phylogenetic groupings within each census region were plotted over 2-month time intervals using Microsoft Excel v16.58.

### Statistical analyses

Spatial statistics were performed in ArcGIS Pro v2.9 using default tools. Epidemiologic comparisons were performed using the biostatistics package RVAideMemoire v0.9-81 [33] in R v4.1.1 [34]. Statistical significance was reported at p<0.05 and indicated in the text, figure, and/or table legends, accordingly.

## RESULTS

### Characteristics of the dataset

From April 27, 2018 to June 9, 2020, the STFCP identified 260 blood culture-confirmed acute typhoid fever cases through a strengthened surveillance system and detected 27 asymptomatic *S*. Typhi fecal shedders through household investigations. We examined case household coordinates for spatial and spatial-temporal clustering, and then incorporated phylogenetic analyses of the sequenced subset of 186 Samoan *S*. Typhi isolates – 172 from acute cases and 14 from asymptomatic shedders – among household contacts (S1 Table). Among the 186 whole genome sequenced isolates, three *S*. Typhi genotypes were identified: genotype 3.5.4 (181/186, 97.3%), genotype 4.1 (4/186, 2.2%), and genotype 3.5.3 (1/186; 0.5%). All but one genotype 3.5.4 isolates could be further categorized into three distinct sub-lineages denoted 3.5.4.1 (n=49), 3.5.4.2 (n=51), and 3.5.4.3 (n=80) and differentiated from all other global *S*. Typhi genomes by single canonical SNPs (S1 Table), as previously described [15]. The sequenced subset of acute cases showed no significant difference in patient characteristics (patient sex, mean age, or age group distribution) when compared to all reported cases (Table 1).

**Table 1.** Comparison of age and sex variables of acute cases associated with an *S*. Typhi whole genome sequence (WGS) versus acute cases missing WGS data

| Variable |  | All cases | Cases, WGS available | Cases, WGS missing | Significance |
| --- | --- | --- | --- | --- | --- |
| Total |  | 260 | 172 | 88 | - |
| Sex | Female, N (%) | 112 (43.1%) | 72 (41.9%) | 40 (45.5%) | p=0.560 |
|  | Male, N (%) | 147 (56.5%) | 99 (57.6%) | 48 (54.5%) |  |
| Age | Mean age (SD) | 24.4 (16.7%) | 24.1 (16.5%) | 25.0 (17.3%) | p=0.709 |
| Age groups | 0-4 years old, N (%) | 21 (8.1%) | 15 (8.7%) | 6 (6.8%) | p=0.864 |
|  | 5-19 years old, N (%) | 103 (39.6%) | 66 (38.4%) | 37 (42.0%) |  |
|  | 20-59 years old, N (%) | 111 (42.7%) | 75 (43.6%) | 36 (40.9%) |  |
|  | 50+ years old, N (%) | 25 (9.6%) | 16 (9.3%) | 9 (10.2%) |  |
| Categorical variables (sex and age group) tested with Pearson's Chi-squared test of independence.<br>Continuous variable (mean age) tested with unpaired two-tailed T test of means.<br>Statistics were performed using the RVAideMemoire v0.9-81 [33] packages in R v4.1.1 [34]. |  |  |  |  |  |

### Spatial clustering and spatial-temporal hotspot analyses of acute cases

When acute case households (n=260) were mapped, areas with many typhoid cases (higher burden) – Apia, northwestern Upolu, southeastern Savaii, and Upolu’s southern coast – were prominent among the four census regions (arrows in Fig 2A). We used the Average Nearest Neighbor (ANN) and Space-Time Hotspot analyses in ArcGIS Pro v2.9 to assess spatial and spatial-temporal clustering, respectively. As expected for a human host-restricted communicable disease, case households (n=260) demonstrated significant clustering by ANN analysis (R_n_<1, p<0.005). However, when grouped by census region (S3A Fig, Table 2), local differences emerged in the degree of clustering, i.e., R_n_ closer to 0, indicating more tightly grouped clusters. Case households in the AUA (n=42), where population density is greatest and where residents receive putatively treated drinking water through a reticulated system, were the least clustered of the census regions (R_n_=0.62). In contrast, case households in the rural ROU (n=55), where districts each have independent (often untreated) water schemes, were the most clustered (R_n_=0.25) (Table 2). Notably, SAV (n=53), which is more rural than ROU, demonstrated a similar degree of clustering (R_n_=0.60) to AUA (Table 2); however, there were ∼8 case households remotely located in the north and west of Savaii far from any nearest neighbor (S3A Fig).

**Table 2.** Results of the Average Nearest Neighbor (ANN) statistic performed on geographic and phylogenetic groupings of *S*. Typhi acute infections.

| Group<sup>a</sup> | No. of points<sup>a</sup> | $\bar{O}$<br>distance<br>(meters) | $\bar{E}$<br>distance<br>(meters) | Nearest<br>Neighbor<br>Ratio ($R_N$) | ANN<br>Z-score<sup>b</sup> | ANN<br>P-value<sup>c</sup> | S3 Fig.<br>Panel |
| --- | --- | --- | --- | --- | --- | --- | --- |
| All cases | 260 | 676.07 | 2312.04 | 0.29 | -21.83 | p<0.005 | A |
| Cases AUA | 42 | 204.87 | 331.34 | 0.62 | -4.73 | p<0.005 | A |
| Cases NWU | 110 | 351.30 | 858.30 | 0.41 | -11.85 | p<0.005 | A |
| Cases ROU | 55 | 607.40 | 2406.22 | 0.25 | -10.61 | p<0.005 | A |
| Cases SAV | 53 | 1800.82 | 3007.99 | 0.60 | -5.59 | p<0.005 | A |
| Cases 3.5.4.1 | 46 | 1589.89 | 3230.86 | 0.49 | -6.59 | p<0.005 | B |
| Cases 3.5.4.2 | 46 | 1211.20 | 2990.57 | 0.41 | -7.64 | p<0.005 | C |
| Cases 3.5.4.3 | 74 | 1710.15 | 3304.99 | 0.52 | -7.94 | p<0.005 | D |
| ANN is a statistical comparison of the observed mean ($\bar{O}$) and expected mean ($\bar{E}$) distances between points yields a nearest neighbor ratio ($R_N$) further described by a probability (p-value) that the observed spatial pattern was created by a random process and standard deviations (z-score). The $R_N$ ranges from 0 (indicating discrete clustering of points) to ~2.15 (indicating uniform dispersion of points equidistant from one another). An $R_N$ value of 1 indicates complete spatial randomness. | | | | | | | |
| <sup>a</sup> Only household coordinates of acute cases were included to assess circulating *S. Typhi* detected through routine passive surveillance and to avoid biasing the point-pattern analyses to households where asymptomatic shedders were localized by active investigation. |  |  |  |  |  |  |  |
| <sup>b</sup> All z-scores are small (<-1.96) corresponding to a confidence level above 95%. |  |  |  |  |  |  |  |
| <sup>c</sup> All p-values are small (<0.005) which indicate statistical significance. |  |  |  |  |  |  |  |

To assess for temporal clustering by census region, a Space-Time Hotspot analysis was performed using a window of 1 km^2^ x 2-months to bin case household coordinates (n=260) and visualized in three-dimensions at two altitudes, 20,000 meters and 10,000 meters, to aid in visualization of each time series (Fig 2B). Temporal persistence of typhoid burden was detectable in AUA as sequential spatial-temporal hotspots, visible as contiguous red columns, (p<0.01) associated with that geographic region (Fig 2B-C). This was in contrast with sporadic (short-lived) hotspots of infection evident in the rest of the country as identified by the discrete red bands (p<0.01) between grey bars in those geographic regions (Fig 2B-C).

### Phylogenetic analysis by genotype, person, place, and time

Subtyping of *S*. Typhi isolates facilitates elucidation of patterns of transmission based on the assumption that similar isolates form a network or chain of infection via a common vehicle and/or source. The presence of three genotype 3.5.4 sub-lineages in Samoa (Fig 3A) allowed the demarcation of useful groupings based on genetic similarity for subsequent, more detailed epidemiologic analyses of associations. The 14 sequenced isolates from asymptomatic shedders detected through household investigations were genotype 3.5.4 and showed no association with any particular sub-lineage (Table 3, Fig 3A). Sub-lineage 3.5.4.1 was significantly (p<0.05) associated with (i) age group (greater-than-expected frequency [GTEF] among ages 0-4 years and 50+; less-than-expected frequency [LTEF] in 5-49 year-olds), (ii) census region (GTEF in ROU, LTEF in AUA and SAV), and (iii) year (GTEF in 2019) (Table 3, Fig 3A). Sub-lineage 3.5.4.2 was significantly (p<0.05) associated with the patient’s sex, being isolated from males more frequently than females (Table 3, Fig 3A). Sub-lineage 3.5.4.3 was significantly (p<0.05) associated with 20-49 year-olds and LTEF in the 5-19 and 50+ year old age groups, as well as census region (common to the four census regions but GTEF in the AUA and ROU) (Table 3, Fig 3A).

**Table 3.** Comparison of epidemiologic variables relating to person, place, and time for each Samoa dominant *S*. Typhi genotype/sub-lineage.

|  | Genotype/Sub-lineage |  |  |  |  |  |  |
| --- | --- | --- | --- | --- | --- | --- | --- |
|  | 3.5.4.1 | 3.5.4.2 | 3.5.4.3 | 4.1 | 3.5.4 | 3.5.3 | TOTAL |
| **Total** | 49 | 51 | 80 | 4 | 1 | 1 | 186 |
| **Status** |  |  |  |  |  |  |  |
| Acute Case | 46 (93.9%) | 46 (90.2%) | 74 (92.5%) | 4 (100%) | 1 (100%) | 1 (100%) | 172 |
| Asymptomatic Shedder | 3 (6.1%) | 5 (9.8%) | 6 (7.5%) | 0 (0%) | 0 (0%) | 0 (0%) | 14 |
| Significance | p=1 | p=0.535 | p=1 | p=1 | p=1 | p=1 |  |
| **Sex** |  |  |  |  |  |  |  |
| Female | 24 (49%) | 16 (31.4%) | 40 (50%) | 1 (25%) | 0 (0%) | 0 (0%) | 81 |
| Male | 25 (51%) | 35 (68.6%) | 40 (50%) | 3 (75%) | 1 (100%) | 1 (100%) | 105 |
| Significance | p=0.404 | **p=0.0470** | p=0.137 | p=0.634 | p=1 | p=1 |  |
| **Age group** |  |  |  |  |  |  |  |
| 0-4 years | 7 (14.3%) | 3 (5.9%) | 4 (5.0%) | 1 (25%) | 0 (0%) | 0 (0%) | 15 |
| 5-19 years | 17 (34.7%) | 23 (45.1%) | 23 (28.8%) | 3 (75%) | 0 (0%) | 1 (100%) | 67 |
| 20-49 years | 15 (30.6%) | 19 (37.3%) | 50 (62.5%) | 0 (0%) | 1 (100%) | 0 (0%) | 85 |
| 50+ years | 10 (20.4%) | 6 (11.8%) | 3 (3.8%) | 0 (0%) | 0 (0%) | 0 (0%) | 19 |
| Significance | **p=0.004** | p=0.363 | **p=0.0004** | p=0.093 | p=1 | p=0.543 |  |
| **Census region** |  |  |  |  |  |  |  |
| AUA | 4 (8.2%) | 7 (14%) | 23 (28.8%) | 0 (0%) | 1 (100%) | 0 (0%) | 35 |
| NWU | 20 (40.8%) | 23 (44%) | 35 (43.8%) | 2 (50%) | 0 (0%) | 1 (100%) | 81 |
| ROU | 22 (44.9%) | 12 (24%) | 13 (16.3%) | 0 (0%) | 0 (0%) | 0 (0%) | 47 |
| SAV | 3 (6.1%) | 9 (18%) | 9 (11.3%) | 2 (50%) | 0 (0%) | 0 (0%) | 23 |
| Significance | **p=0.0013** | p=0.452 | **p=0.0072** | p=0.127 | p=0.312 | p=1 |  |
| **Year** |  |  |  |  |  |  |  |
| 2018 | 15 (30.6%) | 28 (54.9%) | 32 (40.0%) | 1 (25%) | 1 (100%) | 1 (100%) | 78 |
| 2019 | 23 (46.9%) | 20 (39.2%) | 42 (52.5%) | 3 (75%) | 0 (0%) | 0 (0%) | 88 |
| 2020 | 11 (22.4%) | 3 (5.9%) | 6 (7.5%) | 0 (0%) | 0 (0%) | 0 (0%) | 20 |
| Significance | **p=0.0083** | p=0.081 | p=0.328 | p=0.762 | p=0.527 | p=0.527 |  |
| Numbers in parentheses indicate the percentages of the number of times the genotype/sub-lineage was detected for each variable relative to the total number of the genotypes; P-values were calculated using Fisher's exact test using the RVAideMemoire v0.9-81 [33] packages in R v4.1.1 [34]. Bolded p-values are significant at p<0.05. |  |  |  |  |  |  |  |

**Fig 3.**
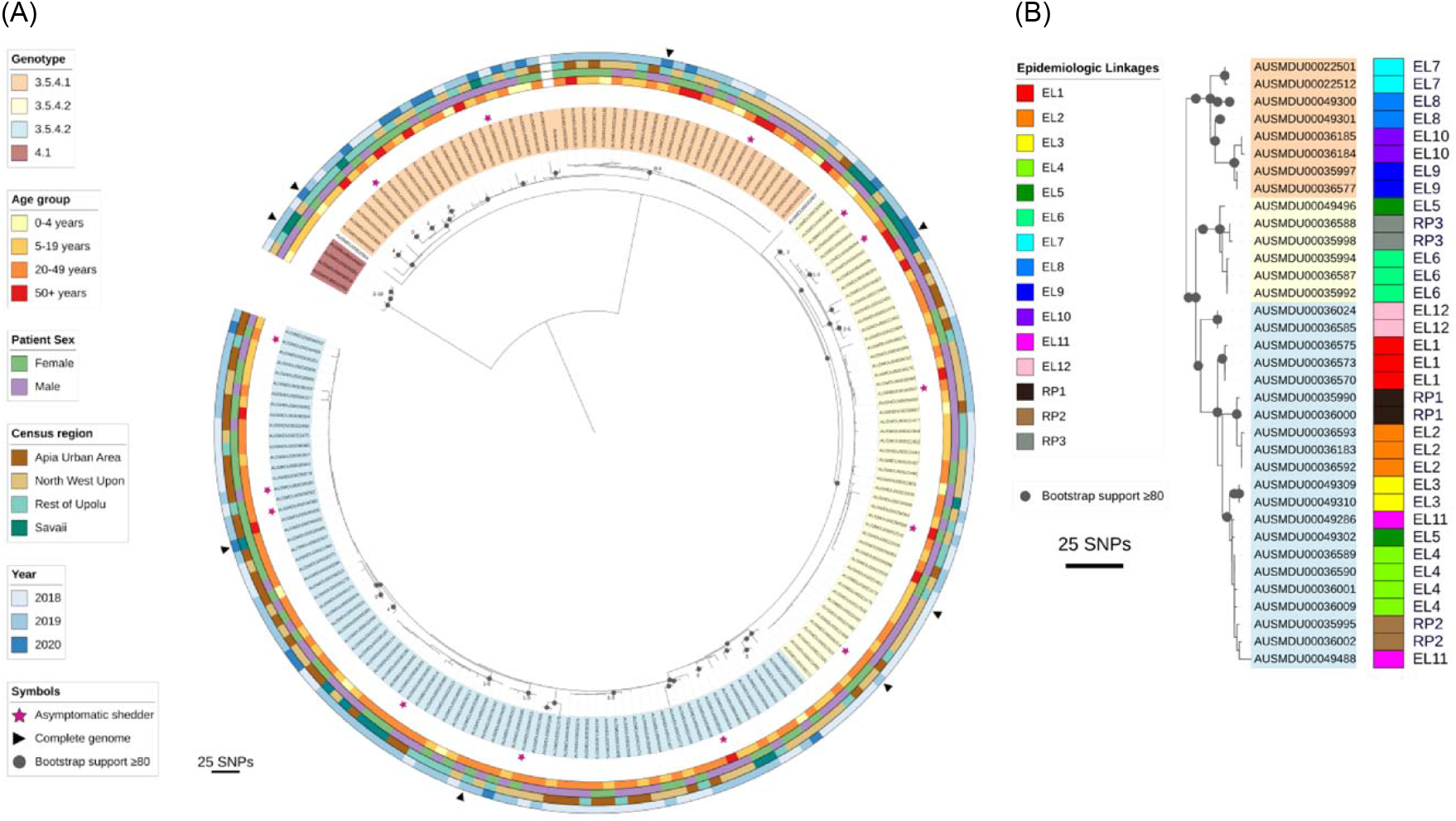
Maximum-likelihood phylogeny, associated epidemiologic variables, and epidemiologic linkages among *S*. Typhi from Samoa from April 2018 through June 2020. **(A)** Maximum-likelihood phylogeny from *de novo* assembled whole genome sequences of Samoan *S*. Typhi genotype 3.5.4 isolates from April 27, 2018 through June 9, 2020 (N=186, S1 Table). Core genome single nucleotide polymorphisms (SNPs) were identified relative to the 2012 Samoa Reference genome H12ESR00394-001A (GCA_001118185.2). Genotype/sub-lineage is indicated by isolate label shading. The labels of the three genotype 3.5.4 sub-lineages 1-3 are identified with orange, yellow, and blue label shading, respectively. Genotype 4.1 isolates are shaded in red. Two outlier isolates are not shaded. Asymptomatic shedders of *S*. Typhi are denoted with a pink star. The patient age group, patient sex, census region of residence, and year of isolation are annotated in the circumscribing colored rings. **(B)** Epidemiologic linkages (EL) identified through case household investigations are compared against genomic relatedness of the isolates based on core genome phylogeny. The visible isolates were extracted from the tree in Panel A. In total 2/12 epidemiologic linkages are not supported by the genomic variation observed in the isolates from those samples (EL5/dark green and EL11/pink groups are not similar). Three repeat positive (RP) blood cultures isolated ∼1 month apart are also compared, showing close genetic similarity.

Geospatially, case households associated with each genotype/sub-lineage were found in the four census regions (Table 3) and were overlapping (Fig 2A), indicating widespread circulation among the islands. Spatial clustering of each 3.5.4 sub-lineage was significant (p<0.005) and intermediate (0.4<R_n_<0.6) among the groupings tested by ANN (Table 2). Within each census region, we identified variation in the frequency as well as transient expansions and contracts in the proportions of each 3.5.4 sub-lineage over time (Fig 4). Total case counts peaked in July-August 2019 and then sharply declined in November-December 2019 and again in March-April 2020 (highlighted background colors in Fig 4), coinciding with three major national events.

**Fig 4.**
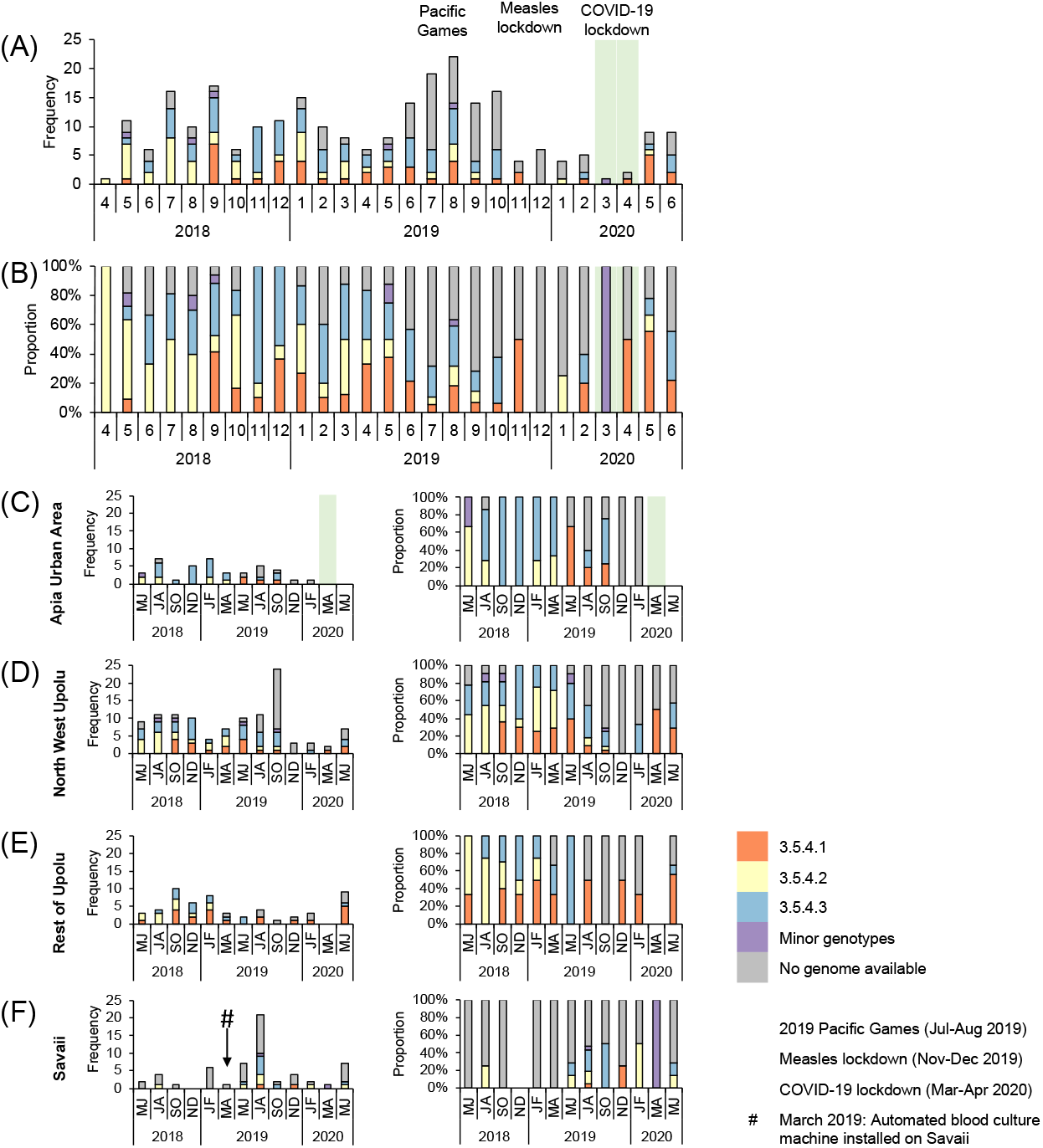
Bar graphs depicting the number and proportion of blood culture-confirmed cases of typhoid fever in Samoa over time from April 27, 2018 through June 9, 2020 by genotype/sub-lineage. **(A-B)** The frequency and proportion of acute cases (N=260) occurring each month are colored by genotype/sub-lineage (n=172) or greyed if no genome was available (n=88). Genotype 3.5.4 sub-lineages 1-3, comprising a total of 166/172 sequenced isolates from acute cases, are identified as orange, yellow, and blue bars, respectively. Minor genotypes (purple bar) include genotype 4.1 (4/186), genotype 3.5.3 (1/186), and one genotype 3.5.4 outlier (1/186) that did not fall into one of the three sub-lineages. **(C-F)** The frequency and proportion of acute cases occurring over 2-month intervals in each census region. Three major events impacting population mobility – increases during the 2019 Pacific Games in Nov-Dec 2019, and restrictions during the measles lockdown from Nov-Dec 2019 [35] and COVID-19 lockdown from Mar-Apr 2020 [36] – are labeled as light red, light purple, and light green background colors, respectively, over the two-month time intervals affected.

### Genetic distances between epidemiologic linkages

WGS analyses can also be used to verify epidemiologic linkages, thereby providing evidence of transmission based on the relatedness of *S*. Typhi isolates. We defined relatedness based on the number of core genome SNP differences between isolates, with fewer differences indicating closer relatedness. During household investigations, the Typhoid SWAT teams attempted to trace infections and link index cases by person, place, and/or time. In 12 instances, genomes were available for putatively linked infections in relation to person, place, and/or time (Fig 3B, Table 4). In six of these, an asymptomatic shedder of *S*. Typhi was identified. In nine epidemiologic linkages, the genotype sub-lineages matched and SNP distances between isolates were two or fewer, indicating close relatedness (Fig 3B, Table 4). In two linkages, EL8 and EL11, the genotype sub-lineages matched, but the SNP distances between isolates were greater, with nine and 14 SNP differences, respectively (Table 2). In another (EL5), the isolates were of completely different *S*. Typhi sub-lineages and 28 SNPs apart (Table 4). Additionally, microbiological surveillance during the study period identified three instances in which two positive *S*. Typhi cultures derived from the same individual were collected ∼1 month apart (Table 4). In each instance, the genotype sub-lineages matched (Fig 3B) and the SNP distances between the first and second isolation were 1-3 SNPs, indicating the isolates were closely related (Table 4).

**Table 4.**
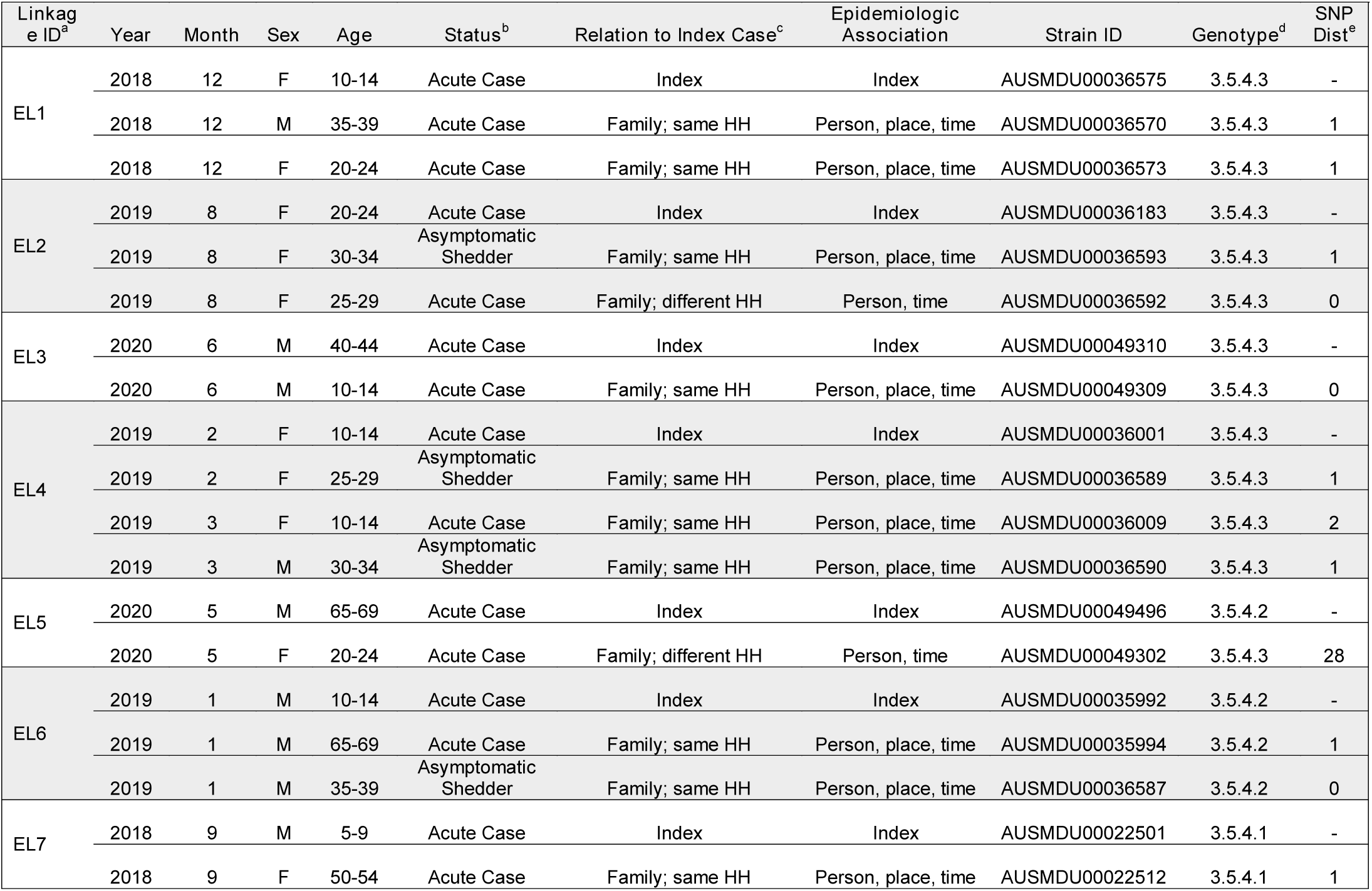
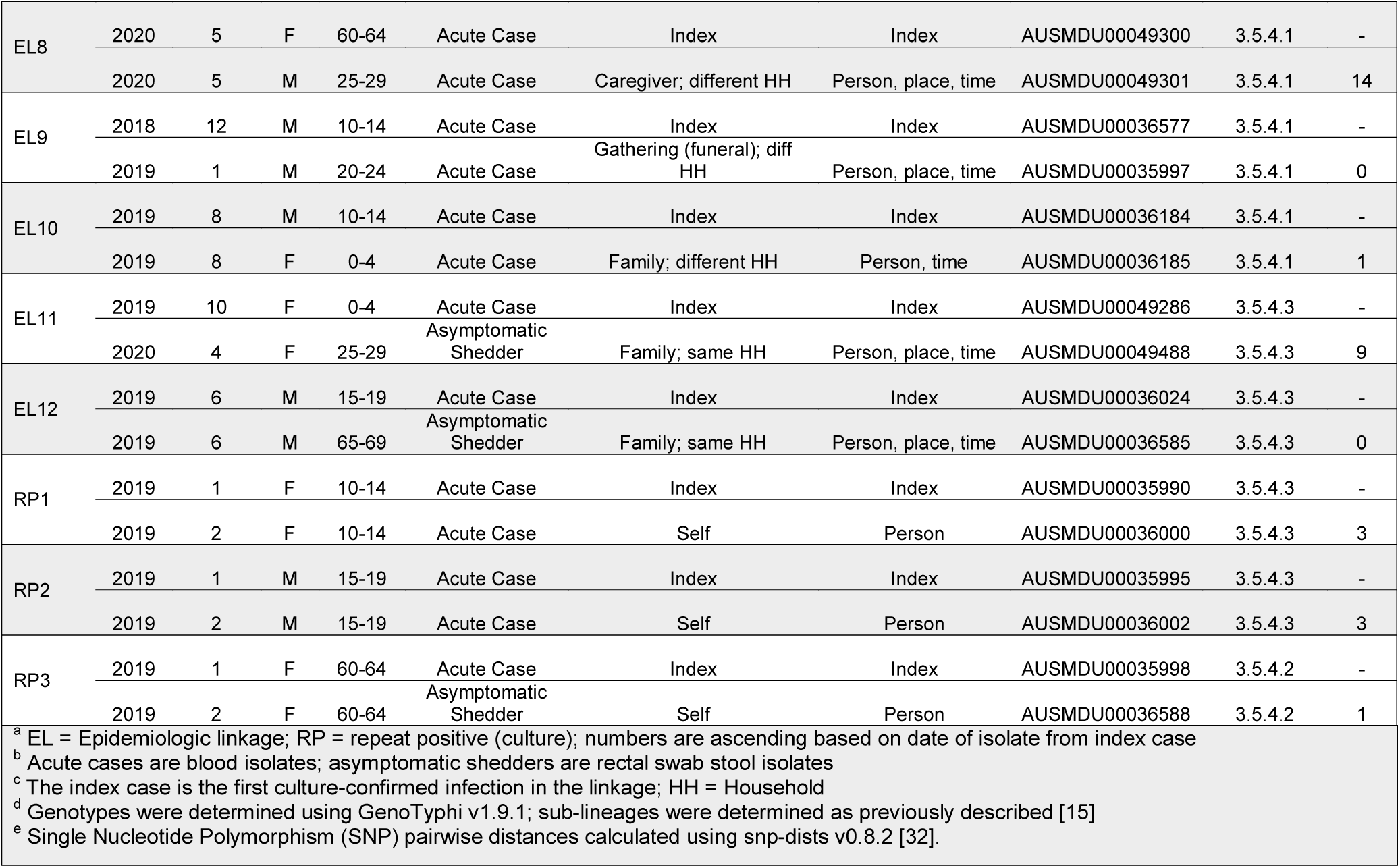
Suspected epidemiologic linkages detected through active surveillance and associated genotype and pairwise SNP distance between isolates

## DISCUSSION

Identifying the modes of transmission that maintain the endemicity of typhoid in Samoa has been a subject of interest for many decades [12,14]. Some insights were derived from our review of the Samoa typhoid burden from 2008-2019 [14]: 53-193 cases of blood culture-confirmed typhoid fever occurred annually without obvious seasonality, equating to a persistently moderate annual incidence. The greatest incidence and numbers of cases was observed among school age children and working age adults in the contiguous regions of NWU and AUA where piped treated water reaches most households, indicating that their daily life patterns put them at enhanced risk. Herein, we relate results of applying the potential discerning power of spatial-temporal cluster and phylogenetic analyses to typhoid infections in Samoa from April 2018 through June 2020.

The burden of typhoid fever was most concentrated in four locations (hotspots; arrows in Fig 2A) determined by point densities: Apia, northwestern Upolu, southeastern Savaii near the ferry terminal, and Upolu’s southern coast. We hypothesized that spatial and temporal differences among the underlying census regions would support either widespread, continuous typhoid circulation or focal, sporadic case clusters around households. Case households in both urban and rural regions showed significant clustering (p<0.005, R_n_<1); however, the distances between cases were much smaller in rural ROU (lower R_n_), indicating stronger foci of spatial clusters compared with more dispersed case clustering (greater R_n_) in AUA (Table 2). In Savaii, the R_n_ was closer to the value for AUA, likely due to skewing from ∼8 individual cases occurring in remote isolation distant from nearest neighbors (S3A Fig). Spatial-temporal hotspot analysis revealed consecutive, significant (p<0.01) temporal hotspots in AUA and the adjacent peri-urban regions of NWU (Fig 4), which could represent a locus of persistent, amplified transmission and also a source of infection for persons from other parts of the islands who work, visit, or consume food prepared in and around Apia.

Contextualization of cases over time amid major national events that either greatly expanded or abruptly halted population mobility highlighted an association between population mobility and case frequency. Typhoid cases, all of which occurred in Samoan people, rose dramatically during the 2019 Pacific Games in July-August 2019 (light red background in Fig 4A) when there was an influx of athletes and supporters. Notably, we did not detect any non-Samoan genotypes during or after this period among the sequenced isolates (Fig 4A), suggesting that dissemination of an imported strain was less likely responsible for the observed increase in cases. Months later, gatherings and population mobility were limited during an intensive nationwide measles lockdown in November-December 2019 (light purple background in Fig 4A) [35], and during COVID-19 lockdowns in March-April 2020 (light green background in Fig 4A) [36]. During these periods of reduced population mobility, the burden of typhoid diminished markedly (Fig 4A). These data highlight a potential role for community interactions and population movement in transmission.

Sub-typing of clinically indistinguishable infections can help to ascertain unique networks of infection. Our previous WGS analyses of Samoan *S*. Typhi from 1983-2020 revealed that dominant Samoa-exclusive genotypes 3.5.4 and 3.5.3 can be divided into sub-lineages based on canonical SNPs [15]. Herein, we analyzed the 2018-2020 *S*. Typhi isolates by sub-lineages in relation to variables of person, place, and time (Table 3). Infections with sub-lineage 3.5.4.1 were more common in ROU and NWU among very young children and older adults who are typically home. Whereas, infections with sub-lineage 3.5.4.3 were more common in urban regions among 20-49 year olds, who represent the dominant workforce [28] that and may be exposed to *S*. Typhi outside the home. Infections with sub-lineage 3.5.4.2 were more common among men than women. These data suggest different groups of people in different regions have variable exposures to *S*. Typhi. Regrettably, because of the disruptions of COVID-19 and reallocation of resources, it was not possible to undertake a case/control study to verify whether adults and school age children who commute from NWU and ROU to work or school in Apia are at significantly increased risk for developing typhoid versus age- and village-matched persons who remain near home.

Spatial clustering by genotype 3.5.4 sub-lineages revealed variable degrees of clustering (Table 2) and considerable overlap among the household clusters where each sub-lineage was detected (Fig 2A), indicating that the sub-lineages are not geographically restricted to any area of Samoa. In fact, fluctuations and lapses in the absolute and relative proportions of the sub-lineages in each census region over time (Fig 4) indicated exchanges among the regions through a mobile agent (either human carrier or food/water vehicle) and/or a heterogeneous reservoir emitting distinct genotypes at different times. The water supplies are not interconnected between regions, making contaminated products or mobile human carriers more likely to be responsible. Evidence to support an important role for carriers are the 27 household contacts of acute cases that were asymptomatically shedding *S*. Typhi. The subset available for WGS analysis (n=14) demonstrated similar phylogenetic distribution of the isolates among each of the three 3.5.4 sub-lineages (Table 3, Fig 3) indicating roughly even contribution to the dominant networks of infection. Our experience suggests that asymptomatic shedders are an important and underestimated reservoir of *S*. Typhi in Samoa, detectable only through active surveillance and detailed case investigations.

We also utilized WGS to verify epidemiologic linkages implicated through the Typhoid SWAT team investigations. Related isolates based on limited genomic SNP differences would support these linkages. In our analysis, nearly all linkages were strongly supported by genetic similarity, with a median pairwise SNP distance of 1 SNP, and demonstrated likely within-household transmission events and contributions by asymptomatic shedders. These findings are consistent with previous typhoid studies in endemic regions where WGS was paired with household investigations in Jakarta, Kathmandu, and Kolkata [37–39] to support the occurrence of within-household transmission events.

There is not a consensus threshold for SNP differences that would associate *S*. Typhi infections in genomic analyses alone. We detected from 0 to 28 pairwise SNP differences among the 12 putative epidemiologic linkages and 3 instances of repeat positive cultures from the same individual collected ∼1 month apart (Table 4). Relatedness criteria of 10 SNPs or less [40] would encompass 13/15 (86.7%) linkages, and 12/13 were within ≤4 SNPs, a suggested cutoff for *Salmonella enterica* [41]. Future studies to characterize *S*. Typhi population diversity in acute infections and asymptomatic carriers by sequencing and comparing multiple colonies obtained from stool cultures collected over time, as has been completed for other enteric pathogens [42], could aid in establishing appropriate thresholds for SNP-based relatedness among *S*. Typhi isolates for cluster identification.

This study has several limitations. Some *S*. Typhi isolates from cases and asymptomatic shedders were unavailable for sequencing, resulting in lost genomic data. Fortunately, all culture-confirmed cases had epidemiologic investigations that included recording the household GPS coordinates and permitting comparisons of the sequenced subset with the total collection of cases; no significant difference in patient sex, mean age, or age groupings was observed (Table 1). Another limitation is that we generally assumed household coordinates were a reasonable indicator of where an individual was infected, a necessary assumption for consistency in the spatial analyses. However, particularly for AUA and NWU cases, there were few epidemiologic investigations of workplaces of adults or schools of schoolchildren, venues where transmission may have occurred.

Sustained surveillance and institution of routine determination of WGS in Samoa could guide help guide interventions during the Consolidation Phase of the STFCP, which will focus on detection of asymptomatic carriers of *S*. Typhi and possible linked acute cases. Following completion of the mass vaccination of all Samoans 1-45 years of age [43], the genotype/sub-lineages of remaining sporadic cases can be compared to pre-vaccination campaign genotypes/sub-lineages. Findings in the current study build upon a functional “Samoan *S*. Typhi framework” that can inform public health decision making and future genomic epidemiology analyses in Samoa as a paradigm for the understudied region of Oceania.

## Data Availability

Raw sequence data have been submitted to GenBank and accession numbers of individual isolates are listed in S1 Table. Assembled genomes are available on Figshare (doi: 10.6084/m9.figshare.18665686).

https://www.ncbi.nlm.nih.gov/bioproject/PRJNA319593

https://doi.org/10.6084/m9.figshare.18665686

## ACKNOWLEDGEMENTS

The authors wish to acknowledge the staff at the Ministry of Health of Samoa for their administrative and technical support of the Samoa Typhoid Fever Control Program, the staff at the Microbiological Diagnostic Unit Public Health Laboratory, Jane Han for her logistical support of the Samoa Typhoid Fever Surveillance Initiative, and Yuanyuan Liang of the Center for Vaccine Development and Global Health for her discussions of statistical methods.

## Supplementary Information

### DETAILED METHODS

Whole genome sequencing, assembly, and mapping

Maximum-likelihood phylogeny

Spatial and temporal point pattern analyses

Average nearest neighbor (ANN) analysis

Space-time hotspot analysis

## SUPPLEMENTARY FIGURES

S1 Fig. Range and interpretation of average nearest neighbor ratio (R_n_) values

S2 Fig. Average nearest neighbor output summary diagram

S3 Fig. Maps of average nearest neighbor datasets

## SUPPLEMENTARY TABLES

S1 Table. Samoa *S*. Typhi isolates analyzed in this study

