## Supplemental Information for "Spatial-temporal and phylogenetic analyses of epidemiologic data to help understand the modes of transmission of endemic typhoid fever in Samoa"

**DETAILED METHODS**

Whole genome sequencing, assembly, and mapping

Maximum-likelihood phylogeny

Spatial and temporal point pattern analyses

Average Nearest Neighbor (ANN) analysis

Space-Time Hotspot analysis

**SUPPLEMENTARY FIGURES**

S1 Fig. Range and interpretation of average nearest neighbor ratio (R_n_) values

S2 Fig. Average nearest neighbor output summary diagram

S3 Fig. Maps of average nearest neighbor datasets

**SUPPLEMENTARY TABLES**

S1 Table. Samoa *S.* Typhi isolates analyzed in this study (separate spreadsheet)

### DETAILED METHODS

#### Whole genome sequencing, assembly, and mapping

In this study, the Microbiological Diagnostic Unit Public Health Laboratory (MDU PHL) received 198 blood isolates from acute cases, of which 26 were excluded from this analysis (12 were duplicates, 13 samples were contaminated, and 1 failed sequencing quality metrics); and 17 isolates from asymptomatic shedders, of which 3 were excluded from analysis (1 was a duplicate; 2 were isolations from food handlers seeking employment registration and thus, were not included in this surveillance study).

Unique dual-indexed DNA libraries were sequenced on the Illumina NextSeq 500 with 150-cycle paired end chemistry. Reads were quality filtered, trimmed, and assembled *de novo* using SPAdes v3.14.1 [1]. Final draft assemblies were filtered to contain only contigs that were ≥500 bp in length and had ≥5x k-mer coverage, as previously described [2]. Complete genomes from this collection were also assembled following the MDU PHL pipeline, as previously described [3]. All raw sequences generated in this study have been deposited on GenBank under BioProject PRJNA319593 under the accessions listed in S1 Table and all assemblies have been deposited on Figshare (doi: 10.6084/m9.figshare.18665686).

Genotypes were assigned with GenoTyphi v1.9.1 [4,5], in which canonical SNPs are determined using the classical *S.* Typhi reference strain CT18 (GCA_000195995.1) as the reference. Samoa-specific sub-lineages (i.e., 3.5.4.1, 3.5.4.2, and 3.5.4.3) were defined as previously described [3].

#### Maximum-likelihood phylogeny

Core genome single nucleotide polymorphisms (SNPs) were determined by alignment to the 2012 Samoan *S.* Typhi reference strain H12ESR00394-001A (GCA_001118185.2) using NASP v1.2.0 [6]. To exclude regions of recombination, the resultant alignment was analyzed with Gubbins v2.4.1 [7]. RAxML v8.2.10 [8] was determined using the PHYLIP format alignment of filtered polymorphic sites using the generalized time-reversible (GTR) site substitution model with a Gamma rate distribution and the Lewis ascertainment bias correction (ASC_GTRGAMMA) and 100 bootstrap pseudo-replicates. Pairwise SNP distances between genomes were calculated using snp-dists v0.8.2 [9]. The resulting ML phylogeny tree was visualized in iTOL v6.5.2 [10], mid-point rooted, and decorated with epidemiologic variables relating to person (sex, age group, host status as a case or asymptomatic shedder), place (census region), time (collection date), and suspected linkages to other infections.

#### Spatial and temporal point pattern analyses

To evaluate the spatial and temporal patterns of circulating *S.* Typhi in Samoa, we performed a series of point-pattern cluster analyses using standard tools in ArcGIS Pro v2.9 [11]. These analyses were focused on the acute cases of typhoid fever investigated by the Samoa Typhoid SWAT teams from 2018 to 2020 in order to assess circulating *S.* Typhi detected through routine surveillance and to avoid biasing the point-pattern analyses to households where asymptomatic shedders were localized by active investigation. GPS coordinates of the households associated with all cases occurring from April 27, 2018 to June 9, 2020 collected during investigations were visualized over thematic objects (administrative boundaries, water bodies, and roads) as a set of OpenStreetMap vector data layers of Samoa purchased from NextGIS, 2020 (https://data.nextgis.com; data license: ODbL). Points were enlarged to dissociate individual households and heatmap densities based on count were inferred to demonstrate hotspots (Fig 2A).

Average Nearest Neighbor (ANN) analysis***.*** The Average Nearest Neighbor (ANN) analysis detects whether or not there is a statistically significant difference between observed mean distance from a point to its nearest neighboring point and the expected mean distance based on hypothetical random distribution [12–15]. ANN can identify structured patterns (i.e., clustering, randomness, or uniform dispersion) for a set of defined points. The null hypothesis is that the points are in complete spatial randomness. First, latitude and longitude coordinates were converted to a local projection coordinate system (American_Samoa_1962_UTM_Zone_2S) to ensure accurate distance measurements between points. ANN was then performed on grouping of the location of the individual with *S.* Typhi isolates based on census regions and phylogenetic relatedness to determine the degree of clustering or dispersion (S3 Fig). At least 30 points are required to generate an accurate nearest neighbor ratio (R_n_) (S1 Fig). Statistical comparison of the observed mean (Ō) and expected mean (Ē) distances between points yields R_n_, which is further described by a probability (p-value) that the observed spatial pattern was created by a random process and standard deviations (z-score). The R_n_ ranges from 0 (indicating clustering of points) to ~2.15 (indicating uniform dispersion of points equidistant from one another) (S1-2 Figs). An R_n_ value of 1 indicates complete spatial randomness (S1-2 Figs).

Space-Time Hotspot analysis. To visualize the distribution and clustering of acute case households in time, we used the space-time hotspot analysis tool implemented in ArcGIS Pro v2.9 [11]. All household coordinates of acute cases were binned into 1 km^2^ x 2-month space-time cubes (x and y dimensions represent space, and the z-axis represents time) (Fig 2C). As there are no uniform selection of spatial and temporal steps, these space-time cube dimensions were empirically optimized according to the size of the islands and the date range spanning 26 months to ensure multiple points could be binned into cubes and 13 time steps measured. The space-time cube was analyzed in the in the Emerging Hot Spot Analysis using default settings. The resulting spatial-temporal hotspots were visualized in three-dimensions over a basemap of Samoa (Fig 2B).

The space time cube aggregated the 260 points representing households of acute cases into 9,216 fishnet grid locations over 13 time step intervals of 2 month each. Each location was a 1 kilometer by 1 kilometer square area. The entire analysis spanned an area 144 kilometers west to east and 64 kilometers north to south. Each of the time step intervals was 2 months in duration so the entire time period covered by the space time cube was 26 months. Of the 9,216 total locations, 120 (1.30%) contain at least one point for at least one time step interval. These 120 locations comprise 1,560 space time bins of which 189 (12.12%) had point counts greater than zero. There was not a statistically significant increase or decrease in point counts over time.

### SUPPLEMENTARY FIGURES

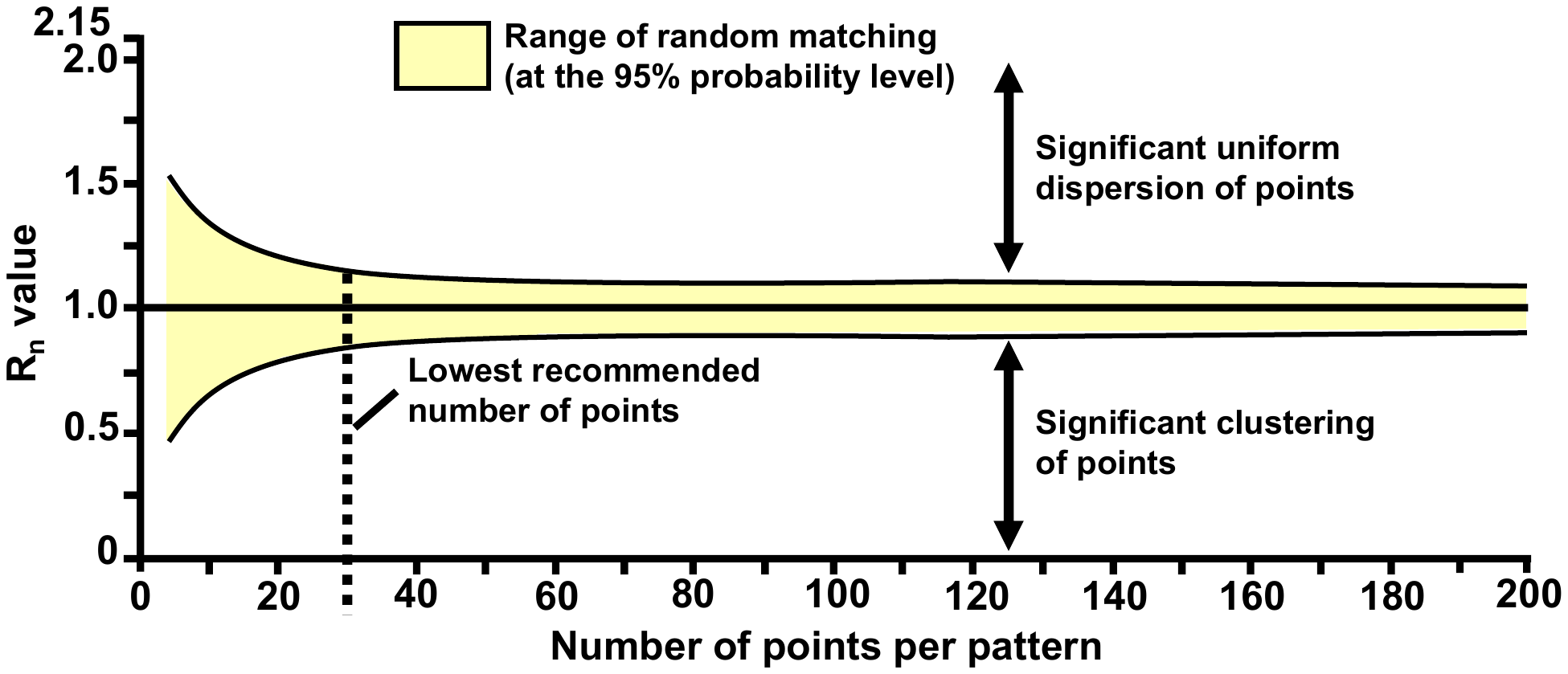

S1 Fig. Range and interpretation of average nearest neighbor ratio (R_n_) values. The R_n_ for a set of points ranges from 0 (clustered) to 1 (random) to ~2.15 (uniformly dispersed). As the number of points per pattern increases, the range of R_n_ values indicating random matching narrows. To approach this narrow range, the minimum recommended number of data points per pattern is 30. This illustration is an adaptation from Pinder et al. (1972), Pinder et al. (1979), and Waugh et al. (2000) [13–15].

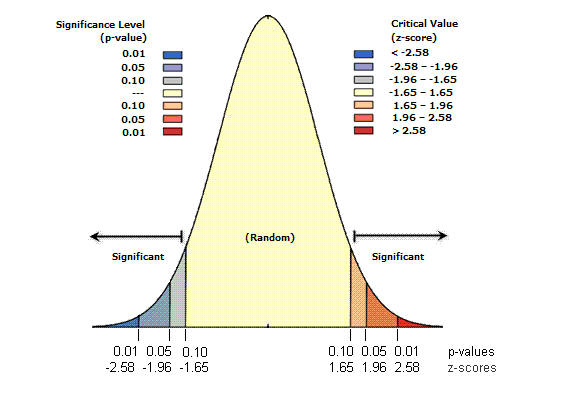

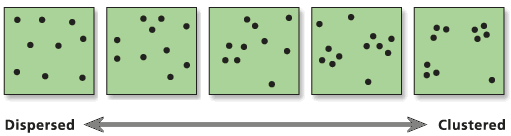

**Uniform**

**R_n_ 🡪 2.15**

**Clustered**

**R_n_ 🡪 0**

**Random**

**R_n_ 🡪 1**

S2 Fig. Average nearest neighbor output summary diagram**.** A visual representation of the nearest neighbor ratio (R_n_) and statistical support values (p-value, and z-score). For example, a set of points with an R_n_ value of 0.5 would indicate clustering. If the z-score (standard deviations) are less than -1.96 or greater than +1.96, then the corresponding p-value will be less than 0.0, and the null hypothesis (i.e., complete spatial randomness) would be rejected with 95% confidence. Adapted with permission from Esri (Redlands, CA: Environmental Systems Research Institute, Inc).

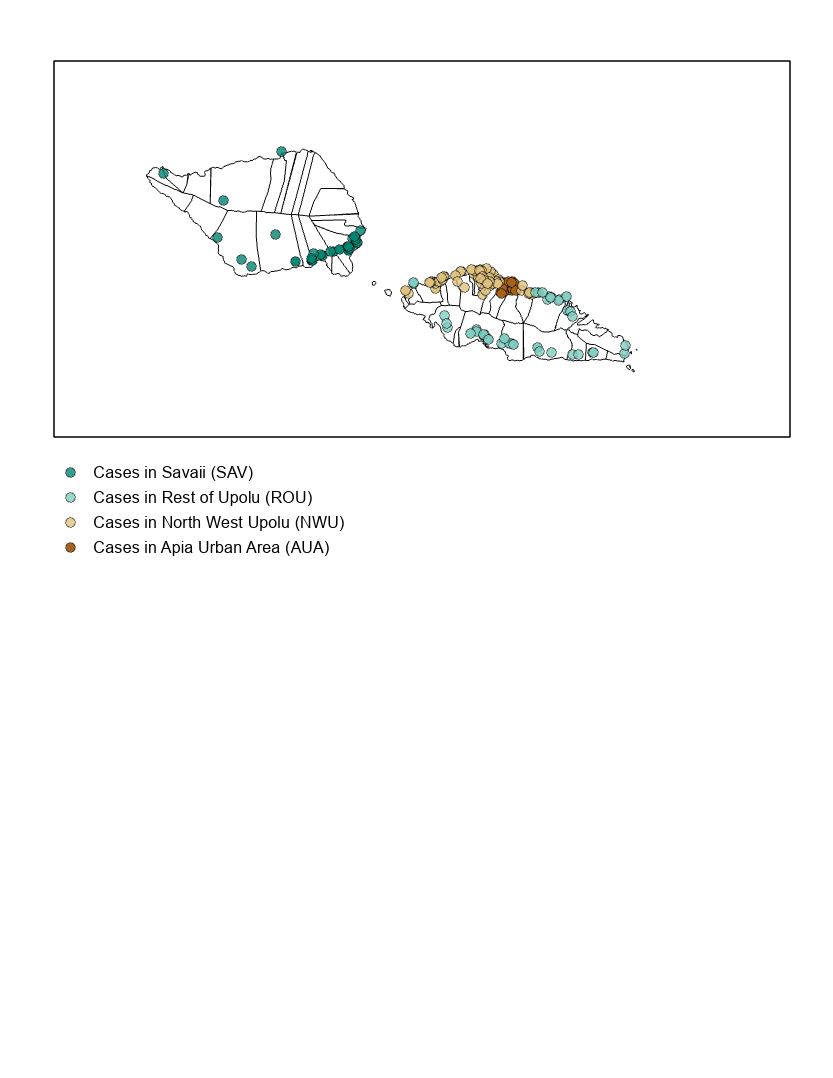

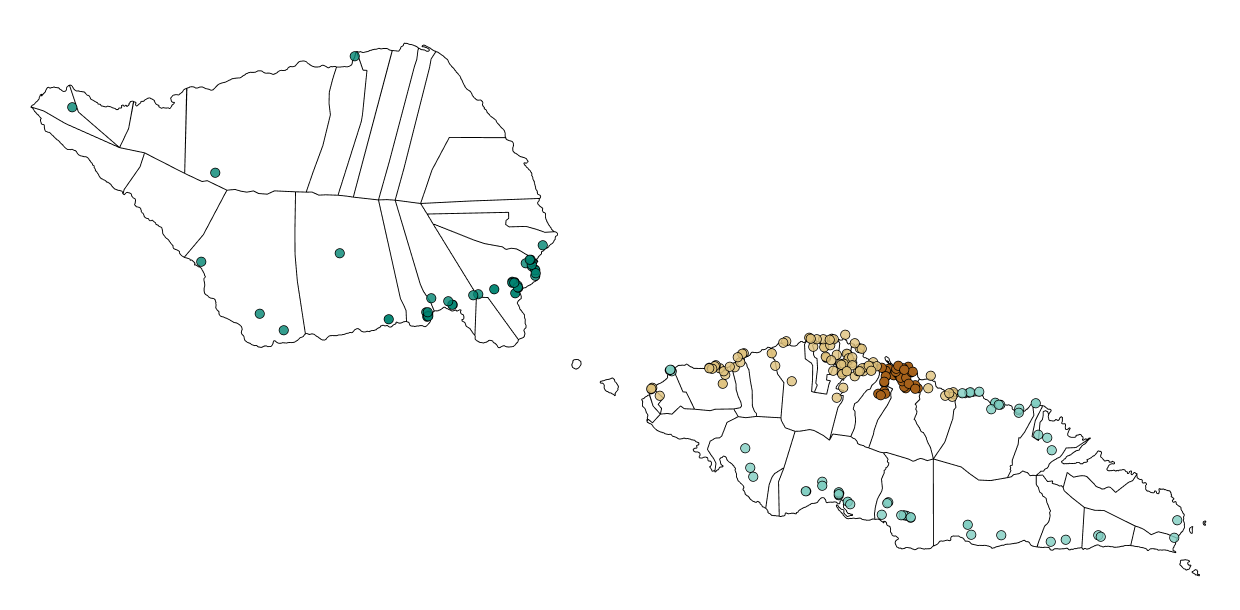

**(A)** All cases (N=260), colored by census region: AUA (n=40), NWU (n=112), ROU (n=55), SAV (n=53)

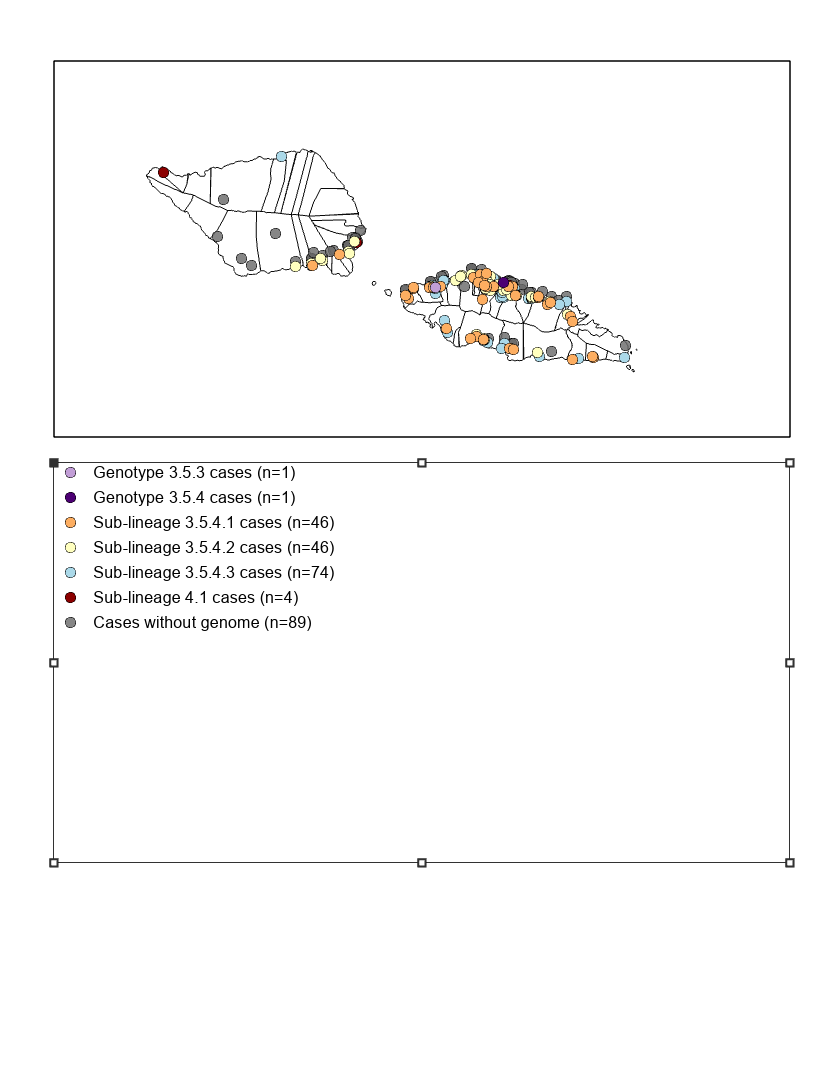

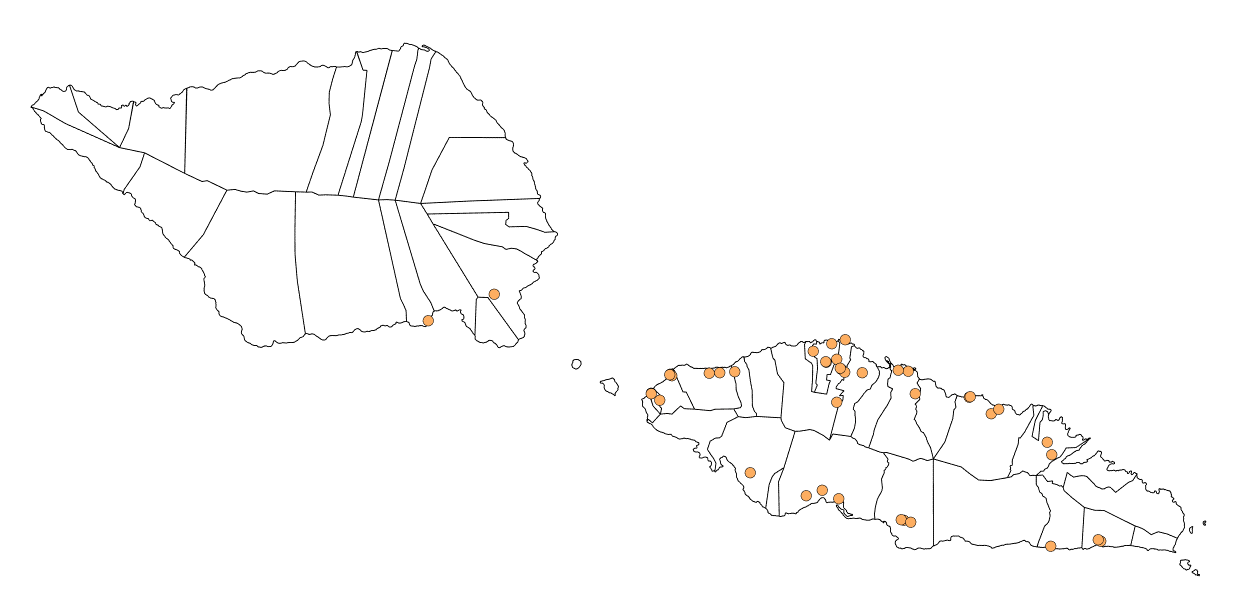
**(B)** All genotype 3.5.4.1 cases

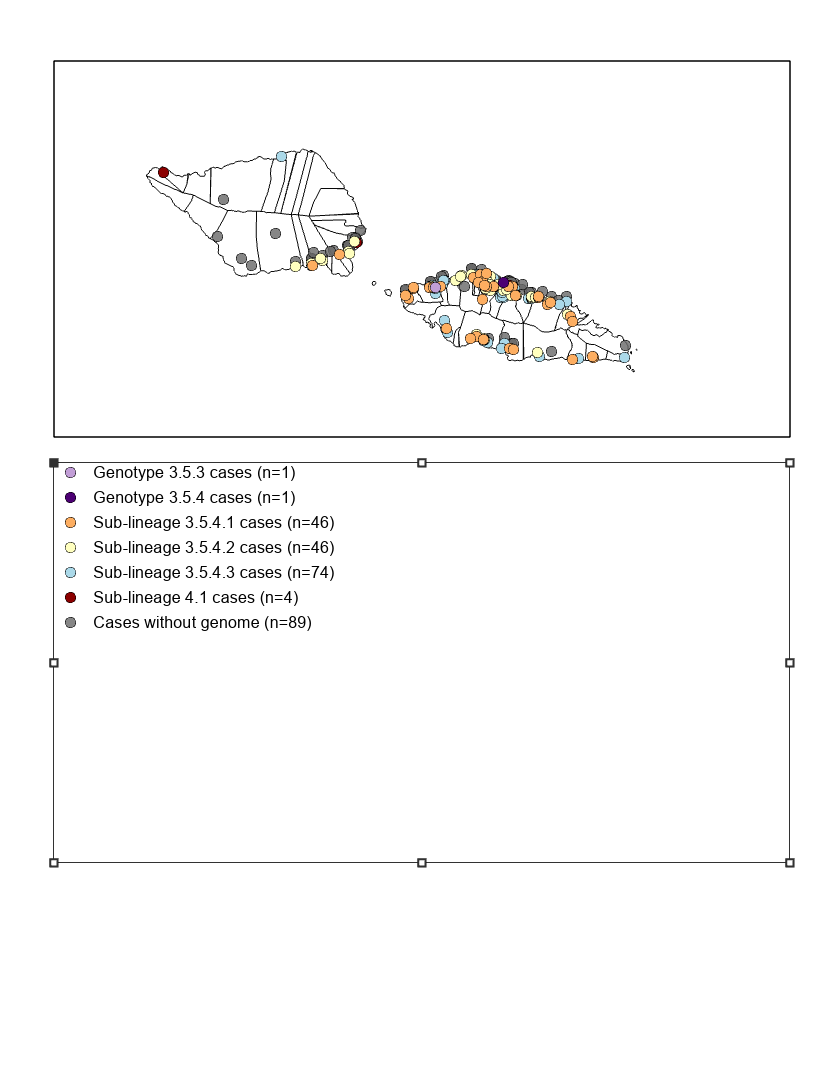

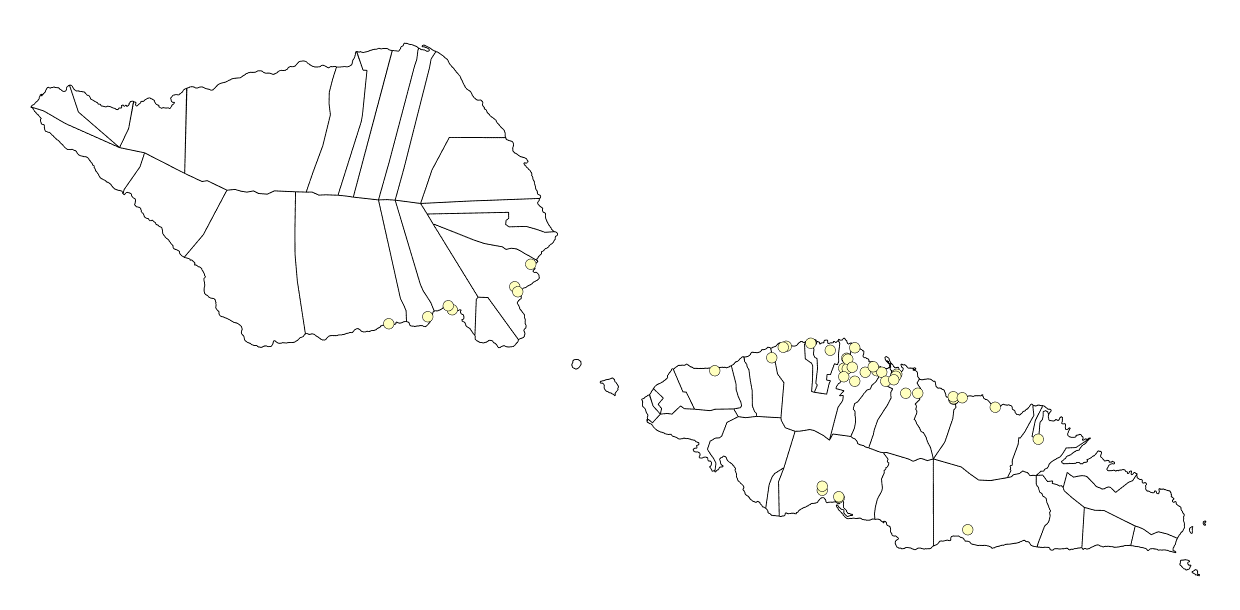

**(C)** All genotype 3.5.4.2 cases

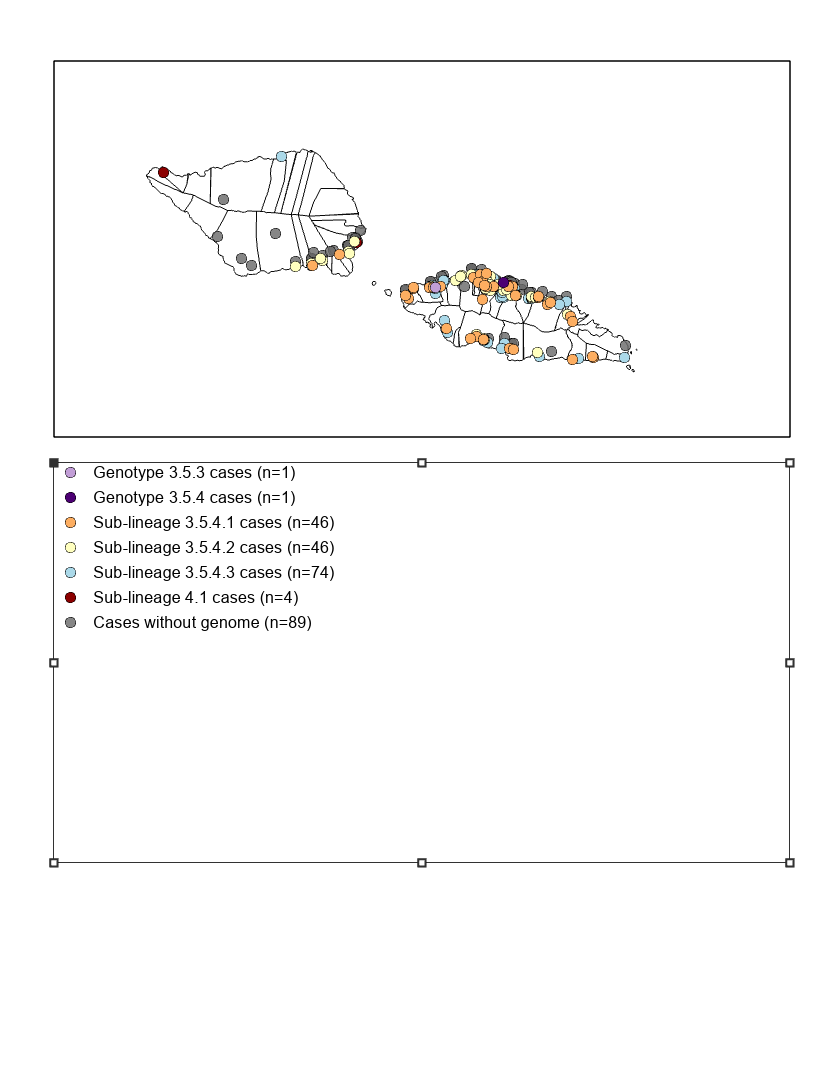

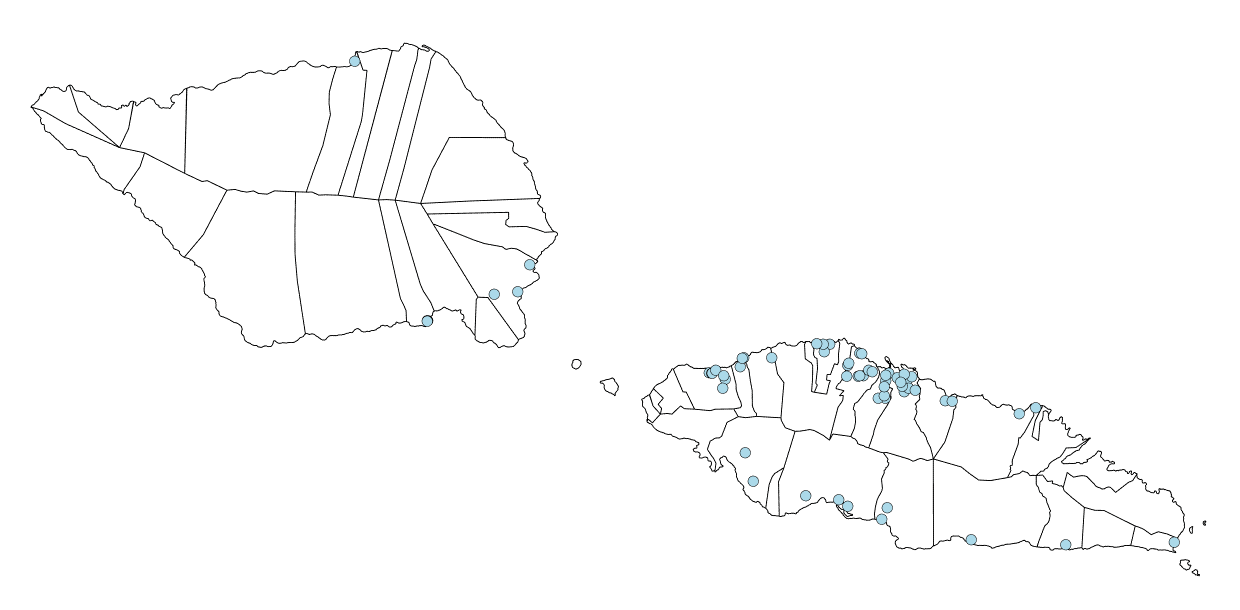

**(D)** All genotype 3.5.4.3 cases

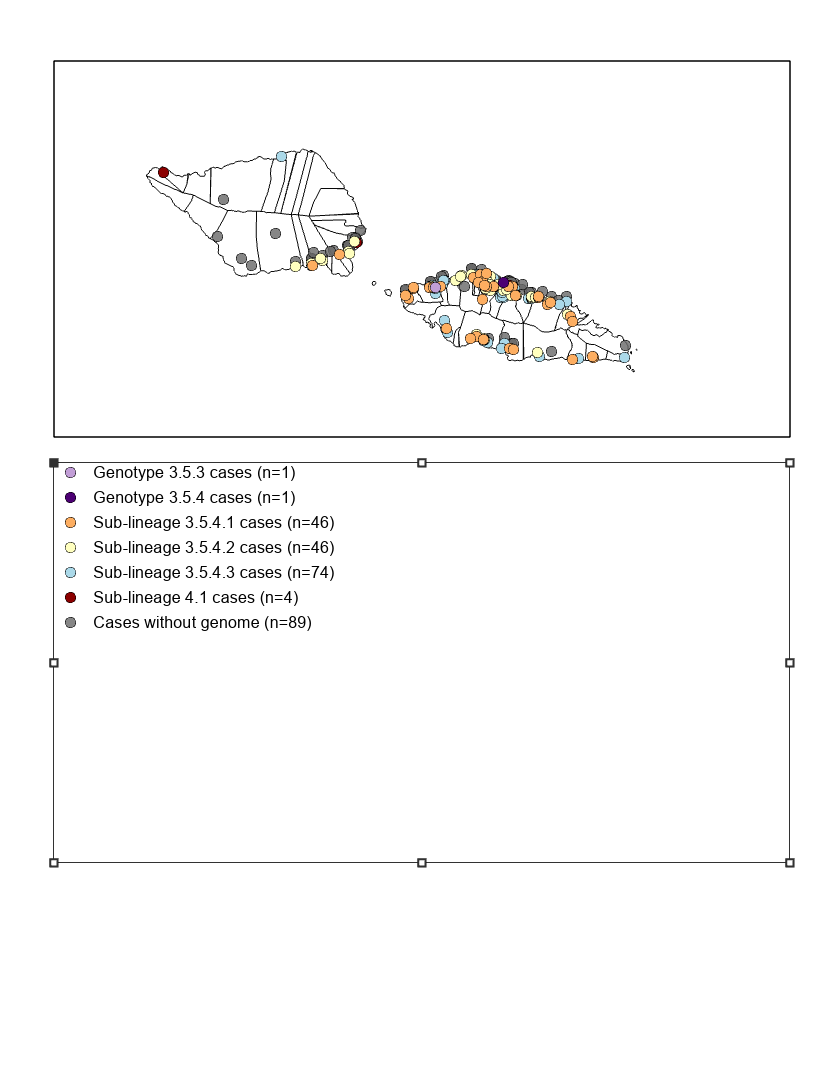

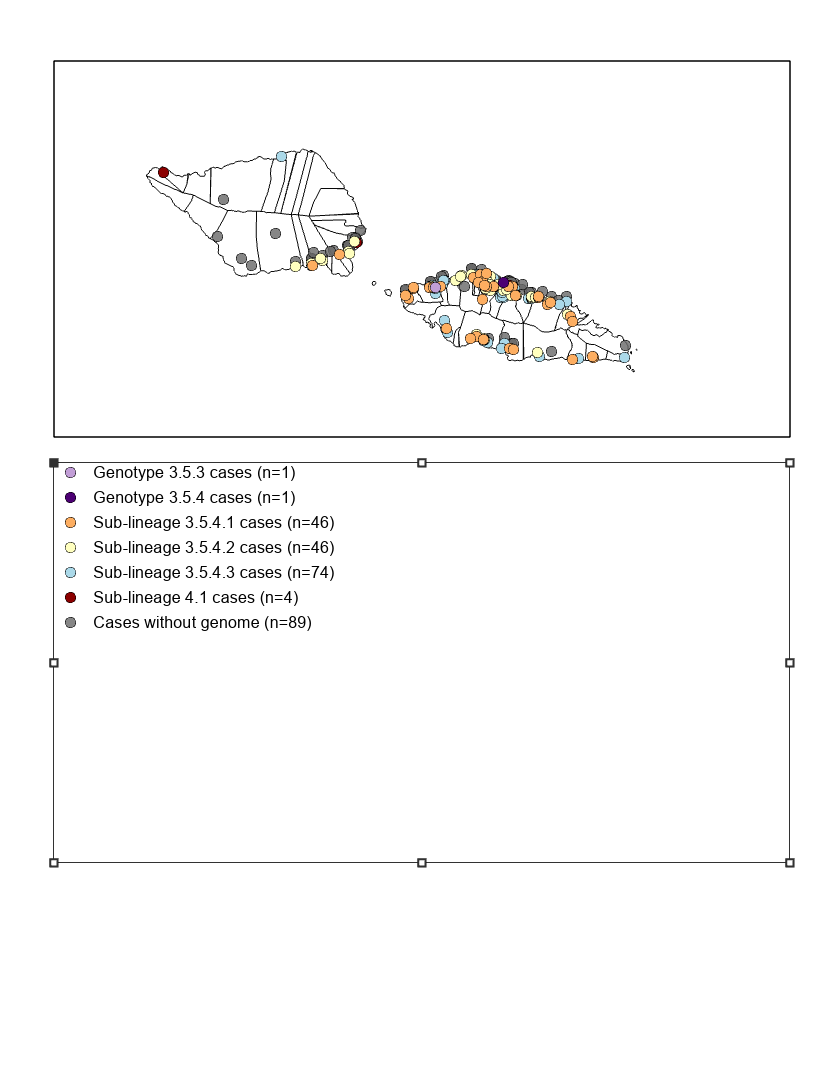

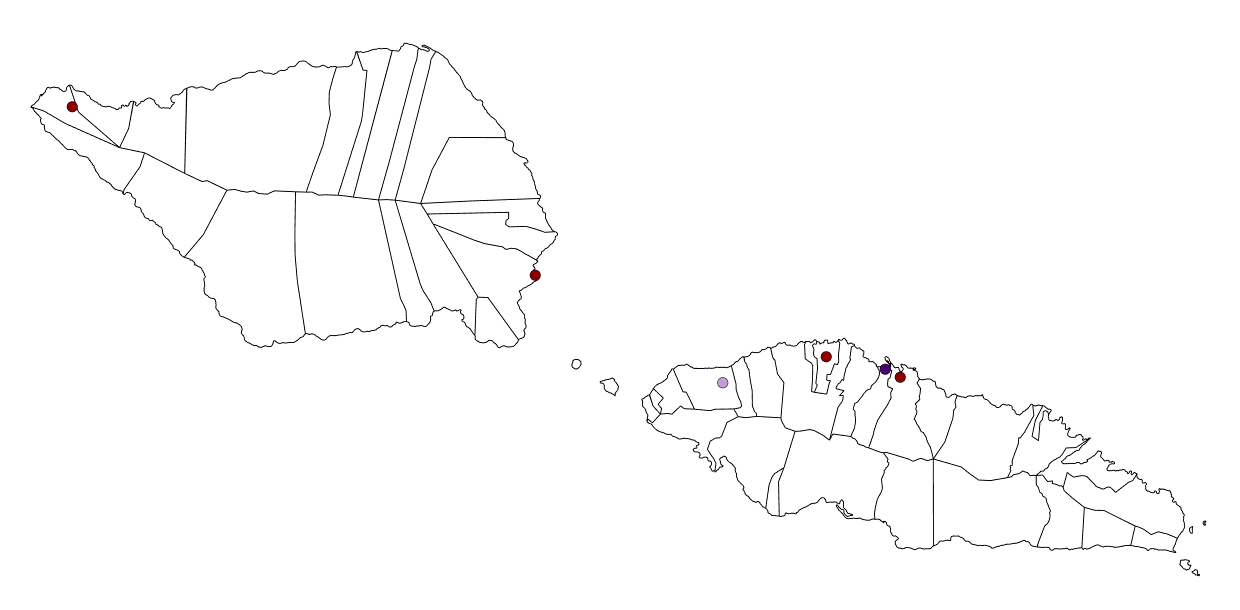

**(E)** All minor genotypes (4.1, 3.5.3, 3.5.4)

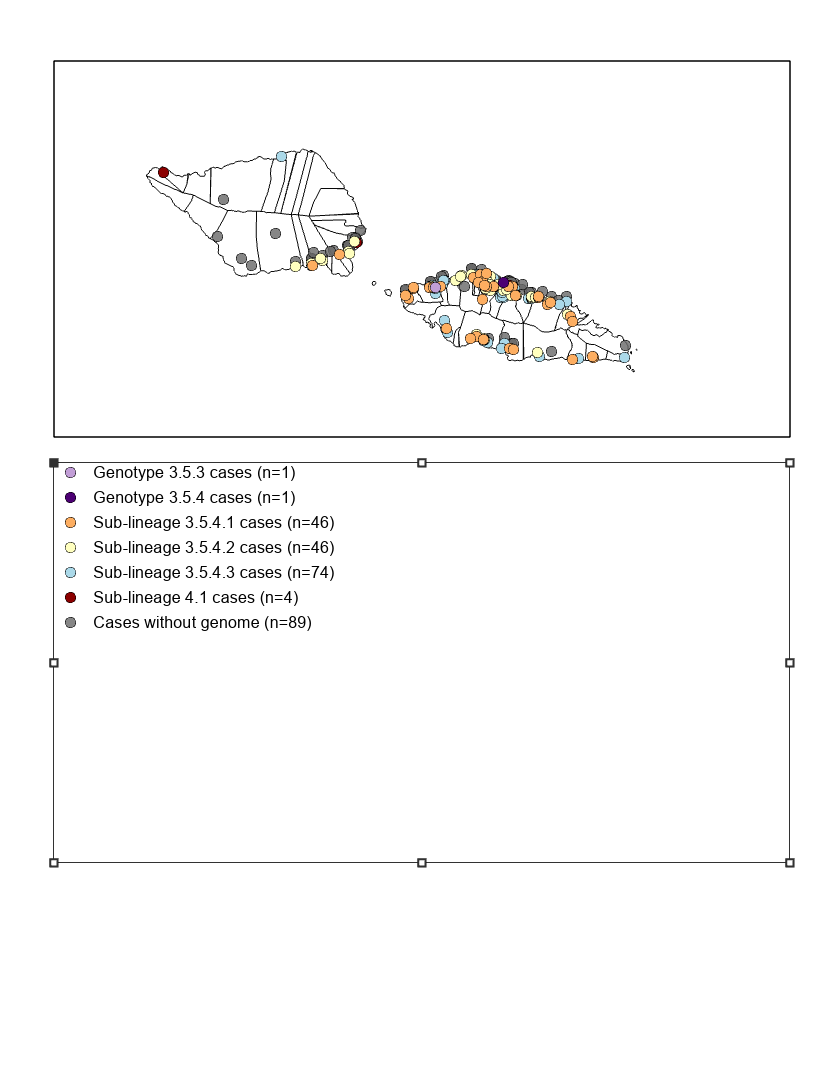

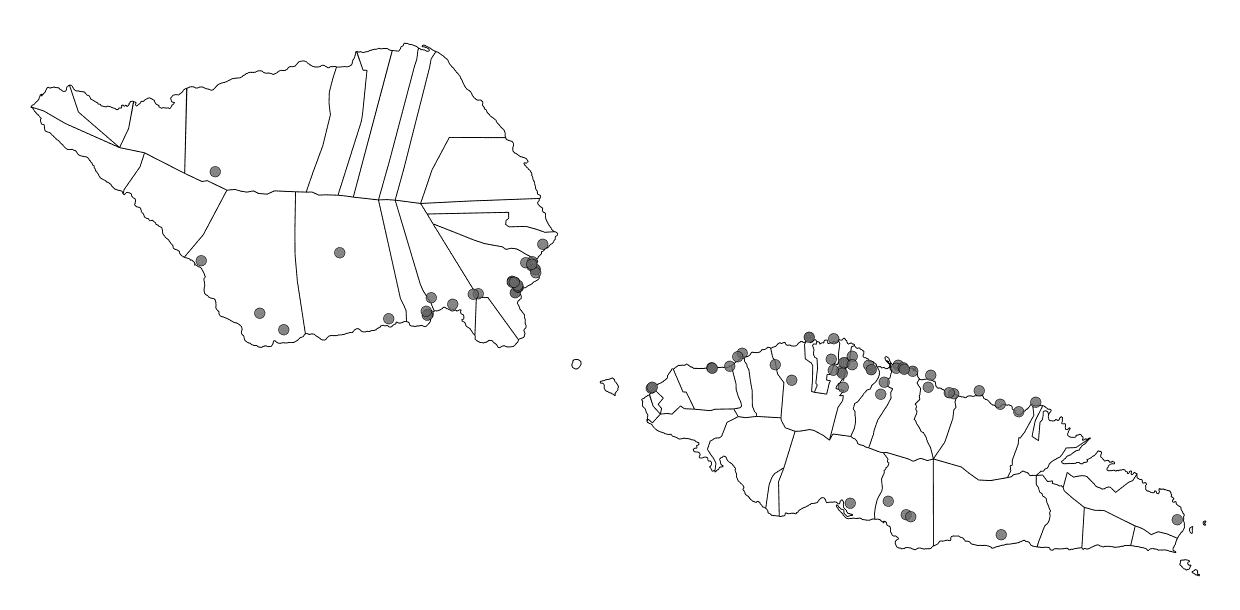

**(F)** All cases without an associated *S.* Typhi genome

S3 Fig. Maps of average nearest neighbor datasets. GPS coordinates corresponding to acute cases of typhoid fever occurring in Samoa from April 27, 2018 through June 9, 2020 grouped by census region and phylogenetic relatedness. (A) All cases colored by census region: Apia Urban Area (AUA) in brown, North West Upolu (NWU) in beige, Rest of Upolu (Rou) in teal, Savaii (SAV) in green. (B) Subset of only genotype 3.5.4.1 isolates in orange. (C) Subset of only genotype 3.5.4.2 isolates in yellow. (D) Subset of only genotype 3.5.4.3 isolates in blue. (E) Subset of only minor genotypes: 4.1 in dark red, 3.5.3 in light purple, and 3.5.4 in dark purple. (F) Subset of cases without an associated *S.* Typhi genome sequence available are in grey.
